# Plasma Metabolomic and Lipidomic Alterations Associated with COVID-19

**DOI:** 10.1101/2020.04.05.20053819

**Authors:** Di Wu, Ting Shu, Xiaobo Yang, Jian-Xin Song, Mingliang Zhang, Chengye Yao, Wen Liu, Muhan Huang, Yuan Yu, Qingyu Yang, Tingju Zhu, Jiqian Xu, Jingfang Mu, Yaxin Wang, Hong Wang, Tang Tang, Yujie Ren, Yongran Wu, Shu-Hai Lin, Yang Qiu, Ding-Yu Zhang, You Shang, Xi Zhou

**Affiliations:** Joint Laboratory of Infectious Diseases and Health, Wuhan Institute of Virology & Wuhan Jinyintan Hospital, Wuhan Institute of Virology, Center for Biosafety Mega-Science, Chinese Academy of Sciences (CAS), Wuhan, Hubei 430023 China; State Key Laboratory of Virology, Wuhan Institute of Virology, Center for Biosafety Mega-Science, CAS, Wuhan, Hubei 430071, China; Center for Translational Medicine, Jinyintan Hospital, Wuhan, Hubei 430023 China; Joint Laboratory of Infectious Diseases and Health, Wuhan Institute of Virology & Wuhan Jinyintan Hospital, Wuhan Jinyintan Hospital, Wuhan, Hubei 430023 China; Department of Critical Care Medicine, Union Hospital, Tongji Medical College, Huazhong University of Science and Technology, Wuhan, Hubei 430030 China; Department of Infectious Diseases, Tongji Hospital, Tongji Medical College, Huazhong University of Science and Technology, Wuhan, Hubei 430030 China; Wuhan Metware Biotechnology Co., Ltd, Wuhan, Hubei 430075 China; Department of Neurology, Union Hospital, Tongji Medical College, Huazhong University of Science and Technology, Wuhan, Hubei 430022 China; State Key Laboratory of Cellular Stress Biology, Innovation Center for Cell Signaling Network, School of Life Sciences, Xiamen University, Xiamen, Fujian 361102 China; University of Chinese Academy of Sciences, Beijing 100049 China

## Abstract

The pandemic of the coronavirus disease 2019 (COVID-19) has become a global public health crisis. The symptoms of COVID-19 range from mild to severe conditions. However, the physiological changes associated with COVID-19 are barely understood. In this study, we performed targeted metabolomic and lipidomic analyses of plasma from a cohort of COVID-19 patients who had experienced different symptoms. We found the metabolite and lipid alterations exhibit apparent correlation with the course of disease in these COVID-19 patients, indicating that the development of COVID-19 affected whole-body metabolism of the patients. In particular, malic acid of the TCA cycle and carbamoyl phosphate of urea cycle reveal the altered energy metabolism and hepatic dysfunction, respectively. It should be noted that carbamoyl phosphate is profoundly down-regulated in fatal patients compared with mild patients. And more importantly, guanosine monophosphate (GMP), which is mediated by not only GMP synthase but also CD39 and CD73, is significant changed between healthy subjects and COVID-19 patients, as well as between the mild and fatal groups. In addition, the dyslipidaemia was observed in COVID-19 patients. Overall, the disturbed metabolic patterns have been found to align with the progress and severity of COVID-19. This work provides valuable knowledge about plasma biomarkers associated with COVID-19 and potential therapeutic targets, as well as important resource for further studies of COVID-19 pathogenesis.

## Introduction

The outbreak of COVID-19, caused by severe acute respiratory syndrome coronavirus 2 (SARS-CoV-2), has been declared a pandemic by the World Health Organization (WHO). Up to the date of April 13, 2020, there are over 1.8 million confirmed COVID-19 cases and more than 110,000 deaths worldwide according to the situation report of WHO. Based on a recent study of 44,672 confirmed COVID-19 cases up to February 11 by Chinese Center for Disease Control and Prevention, over 19% COVID-19 patients developed severe or critical conditions (1). The global fatality rate is around 4.8% in all the confirmed cases until March 31, and has even reached 10% in some developed countries probably due to a more elderly population (2).

The main attacking organ of SARS-CoV-2 is low respiratory tract, and some patients develop life-threatening acute respiratory distress syndrome (ARDS). Besides, the attacks of liver, muscle, gastrointestinal tract, lymph node, and heart by SARS-CoV-2 have also been found or proposed (3-6). On the other hand, although more than 80% COVID-19 patients experienced only mild symptoms, it has been found that the conditions can rapidly progress from mild to severe ones, particularly in the absence of adequate medical care. Moreover, the mortality rate of COVID-19 in critically ill cases can be over 60%, posing great pressure on treatment (7). However, the physiological changes associated with COVID-19 under different symptomatic conditions are barely understood.

Metabolites and lipids are major molecular constituents in human plasma. During critical illness, metabolic and lipid abnormalities are commonly observed, which are believed to contribute to physiology and pathology. Moreover, previous studies have demonstrated dramatic alterations of metabolome and lipidome in human plasma caused by various diseases including viral infections, like Ebola virus disease (8, 9). Here, we performed the targeted metabolomic and lipidomic profilings of plasma samples collected from a cohort of COVID-19 patients, including COVID-19 fatalities and survivors recovered from mild or severe symptoms. Our findings here show many of the metabolite and lipid alterations, particularly ones associated with hepatic functions, align with the progress and severity of the disease, which would provide valuable knowledge about plasma biomarkers associated with COVID-19 as well as potential therapeutic targets, and shed light on the pathogenesis of COVID-19.

## Results

### Study design and patients

Blood samples were harvested at Wuhan Jinyintan Hospital from COVID-19 patients who were confirmed by laboratory nucleic acid test of SARS-CoV-2 infection. Serial samples were collected over the course of disease from 9 patients with fatal (F) outcome (F1-F4), 11 patients diagnosed as severe (S) symptoms (S1-S2), and 14 patients diagnosed as mild (M) symptoms (M1-M2) (Table S1). Of note, all the patients in the severe (S) and mild (M) groups had survived from COVID-19 and been discharged from the hospital. F1 represents the first samples collected from the COVID-19 fatal patients, while F4 represents the last samples before additional samples could be collected. S1 or M1 represents the samples during the disease peak of the patients in the severe or mild group as being determined based on the Diagnosis and Treatment Protocol for Novel Coronavirus Pneumonia (6th edition) published by the National Health Commission of China (10), while S2 or M2 represents the last samples collected from patients in each group before the patients discharged from the hospital. For comparison, the blood samples from 10 healthy volunteers, whose throat swabs and serological testing were negative for SARS-CoV-2, were collected. The hydrophilic and hydrophobic metabolites were extracted from each plasma sample, respectively and measured by employing liquid chromatography electrospray ionization tandem mass spectrometry (LC-ESI-MS/MS) system. The metabolite identification was conducted by home-made database with retention time and ion pairs. For those metabolites without authentic standards in our database, we still used MS/MS spectra to search against the public databases for improving the confidence of metabolite identification. The orthogonal partial least-squares discriminant analysis (OPLS-DA) was used to discriminate metabolomics profiles between the groups of COVID-19 patients and healthy people (Figure S1-S4). In total, 431 metabolites and 698 lipids were identified and quantified, and both metabolome and lipidome showed dramatic alterations in the plasma of these COVID-19 patients (Table S2 and S3).

### Plasma metabolomic alternations associated with clinical symptoms of COVID-19

For different courses of fatal COVID-19 patients (F1-F4), we analyzed the metabolites that underwent significant change [F4 vs. H, >1 log_2_ fold change (FC) <-1, typically *P* <0.05]. For F vs. H, 87 of the total 431 metabolites were significantly different (*P* < 0.05) at F1, while the number of significantly altered metabolites were increased to 162 at F4 in the fatalities; and most of the changes are down-regulated (Table S2). We found a positive correlation between the alteration of metabolites and the course of disease deterioration in fatal patients (Figure 1A and Table S4), indicating that the development of disease affects the metabolism of metabolites.

**Figure 1.**
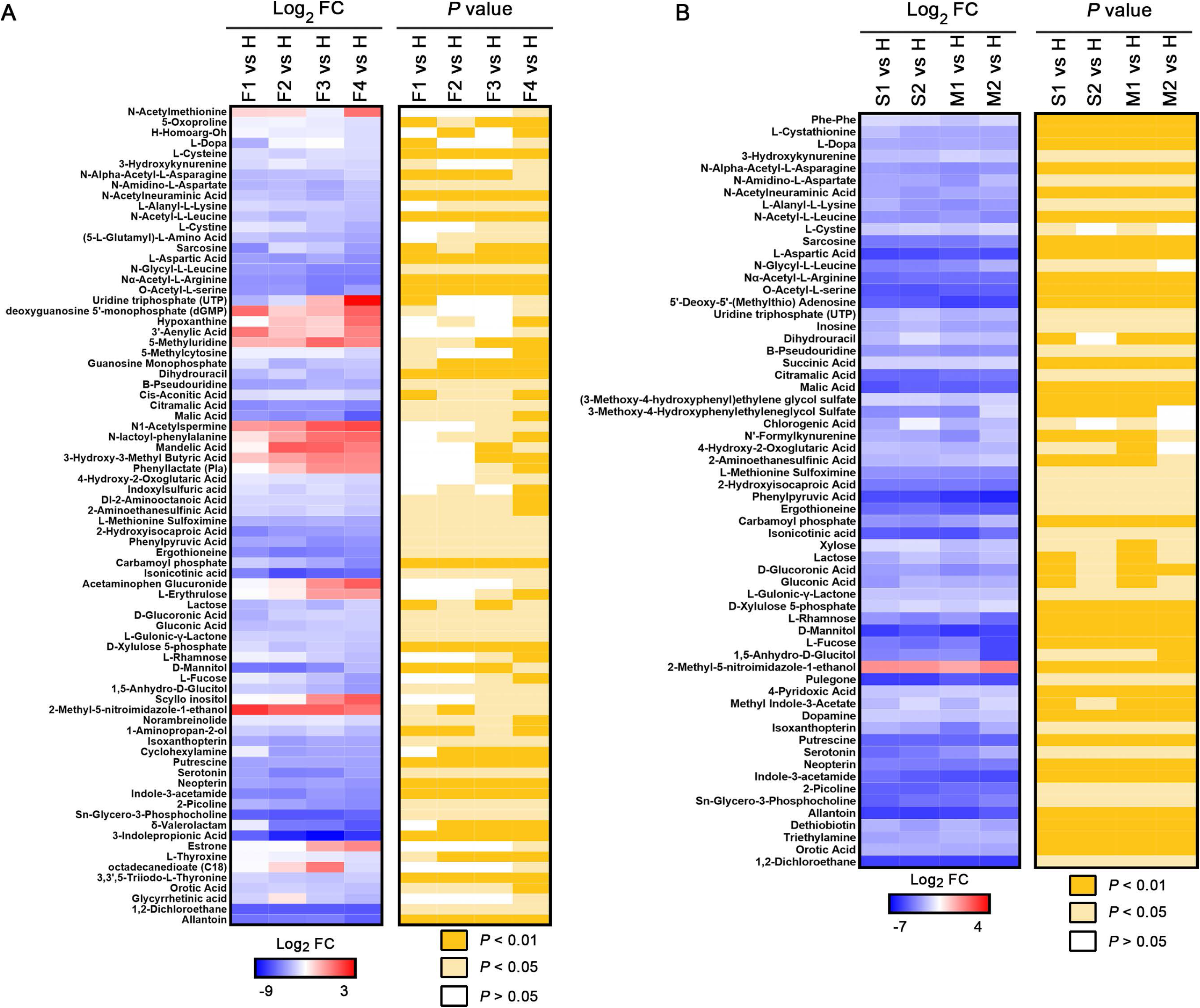
COVID-19 signatures in the plasma metabolome. Selected average plasma metabolite expression levels and associated *p* values for COVID-19 fatality patient group vs. healthy volunteer group (A), and severe or mild vs. healthy groups (B). F, fatalities, first, second, third and fourth samples, F1, F2, F3 and F4. S, severe patients, first and second samples, S1 and S2; M, mild patients, first and second samples, M1 and M2.

We also profiled the metabolites in the different courses of severe and mild COVID-19 patients (S1 and S2; M1 and M2), and analyzed those that underwent the significant change [S1 vs. H, >1 log_2_ FC <-1, typically *P* <0.05; M1 vs. H, >1 log_2_ FC <-1, typically *P* <0.05] (Figure 1B and Table S5). There are apparently less metabolites with significant changes (>1 log_2_ FC <-1, typically *P* <0.05) observed in severe and mild patient groups when compared with those of fatal patients, and almost all the significantly altered metabolites were down-regulated. These results indicate that the alterations of metabolic pathways were more dramatic in fatal COVID-19 cases than in severe and mild ones who finally survived.

In addition, it is noteworthy that although the patients in both severe and mild groups had met the hospital discharge criteria in the time points S2 and M2 as their COVID-19 nucleic acid tests for turning out to be negative twice consecutively, our metabolomic data clearly show that many of their metabolites had not returned to normal levels when compared with those in healthy volunteers (Figure 1B), suggesting that these discharged patients had not been fully recovered from the impacts of COVID-19 in physiology.

To further analyze the metabolomic data, the differentiating metabolites were divided into those shared by all groups (F vs. H, S vs. H, and M vs. H) or those unique to the fatal group (F vs. H). Then, we performed Kyoto Encyclopedia of Genes and Genomes (KEGG) functional enrichment analysis to annotate the potential functional implication of differentiating metabolites among these groups (Figure 2). As shared by all the three symptomatic groups, differentiating metabolites were enriched in total 12 pathways and significantly enriched in 3 pathways including pyrimidine metabolism, fructose and mannose metabolism, and carbon metabolism (Figure 2A-B and Table S6). On the other hand, in the case of the fatality group, differentiating metabolites were significantly enriched in 4 pathways, including thyroid hormone synthesis, thyroid hormone signaling, purine metabolism, and autoimmune thyroid (Figure 2C-D and Table S7), suggesting that the alterations in these pathways are associated with the progress and deterioration of COVID-19.

**Figure 2.**
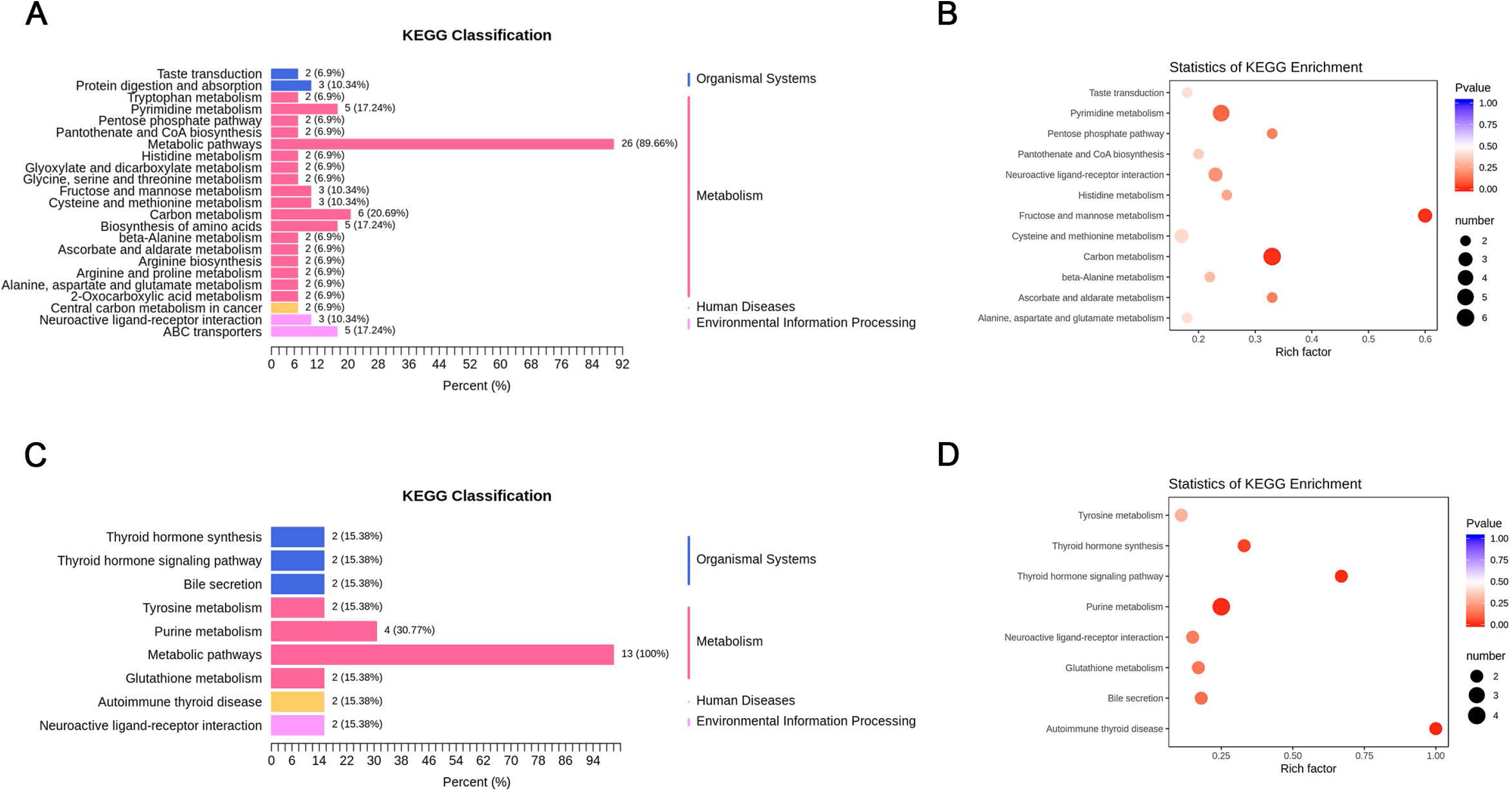
The metabolome KEGG enrichment analysis of COVID-19 patient plasma. (A-B) KEGG pathway analysis of DEMs shared in all the groups. The color of bubbles represents the value of adjusted *P* value, and the size of bubbles represents the number of counts (sorted by gene ratio). (C-D) KEGG pathway analysis of DEMs shared unique to the fatal groups.

A prominent signature observed among fatal COVID-19 patients was an acute reduction of metabolites in patient plasma with the aggravation of the course of COVID-19. By comparing healthy subjects and fatal patients, we highlighted top 5 differentiating metabolites (Figure 3). For instance, malic acid, an intermediate of the tricarboxylic acid (TCA) cycle, exhibited the greatest log_2_ FC (−5.2) among all significantly altered metabolites in the fatalities. Similarly, aspartic acid shows markedly down-regulated in the plasma of patients (Table S4 and Table S5). Thereby, we postulate that a deficiency of malic acid as well as dysfunction of malate-aspartate shuttle may reveal energy depletion and physical exhaustion in COVID-19 patients. These biomarkers are rapidly consumed in inflammatory states to provide energy and materials for the proliferation and phagocytosis of immune cells (11), indicating that immune system were activated in these cases.

**Figure 3.**
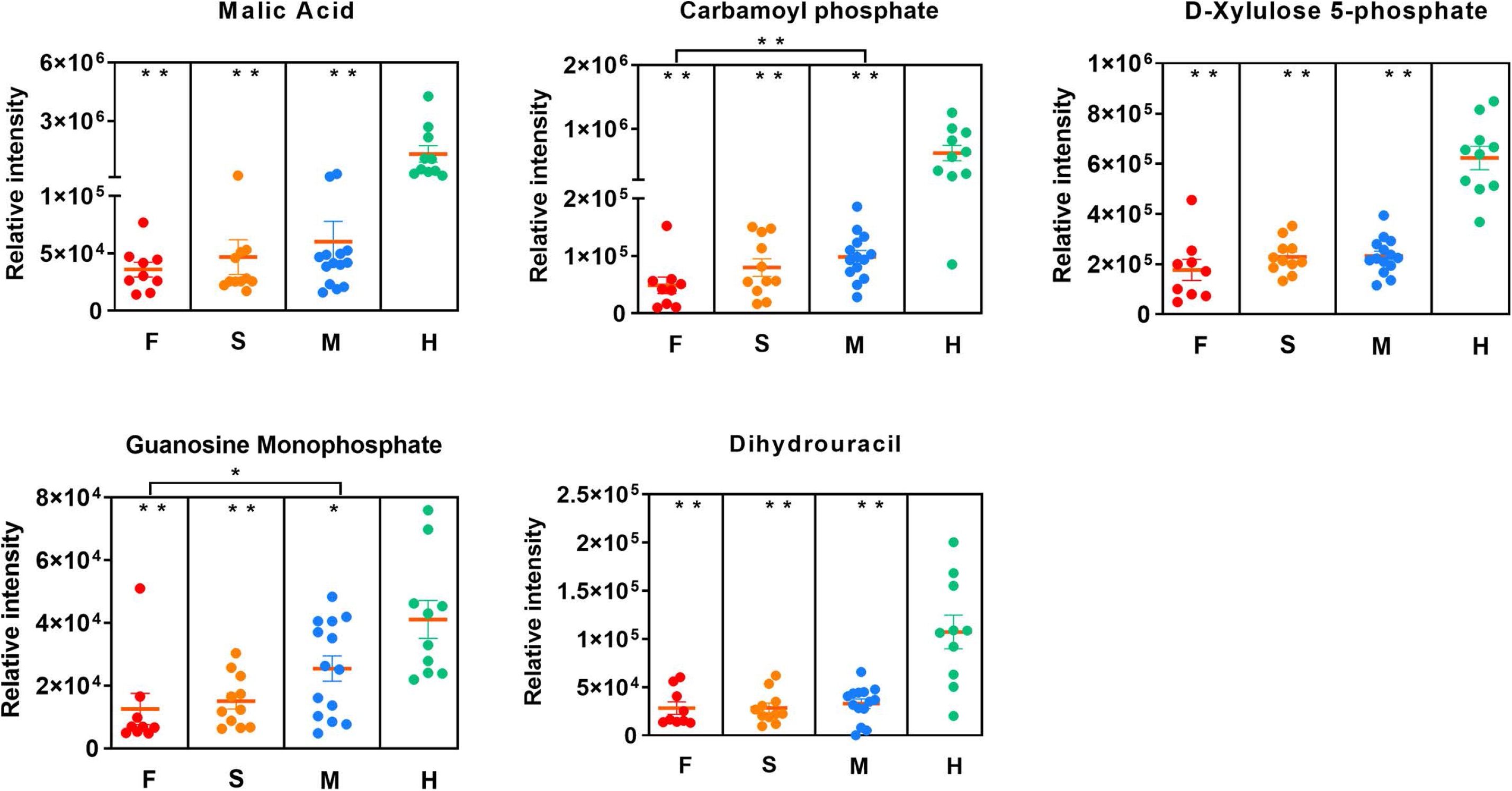
The potential metabolomic biomarkers of COVID-19. The relative intensity for each metabolite. Each dot represents a patient sample, and each patient group is differently colored as indicated. F, fatalities; S, the patients with severe symptom; M, the patients with mild symptom; H, healthy volunteers. \**P*<0.05, \*\**P*<0.01.

Intriguingly, we observed that the levels of some nucleotides and organic acids were significantly increased (e.g., hypoxanthine), whereas the levels of some nucleotides and organic acids were significantly reduced [e.g., guanosine monophosphate (GMP)]. Hypoxanthine-guanine phosphoribosyl transferase (HPRT) is an important enzyme involved in nucleotide recycle pathway and can covert hypoxanthine and guanine to inosine 5’-monophosphate (IMP) and GMP, respectively (12, 13). The observed abnormal levels of hypoxanthine and GMP suggested that the function of HPRT had become defective in these COVID-19 fatal patients, which could result in the disorders of purine and pyrimidine metabolism. Furthermore, GMP is also involved the metabolic reactions mediated by not only GMP synthase but also CD39 and CD73. Both CD39 and CD73 are the immunomodulatory enzymes, suggesting that CD39/CD73 axis imbalance may occur in COVID-19 patients. In addition, we observed that the level of carbamoyl phosphate was significantly and gradually reduced over the course of COVID-19 fatalities. Carbamoyl phosphate is synthesized from free amino donors by carbamoyl phosphate synthetase I (CPSI) in mitochondria of liver cells, and participates in the urea cycle to remove excess ammonia and produce urea (14-17). Its reduction in fatal cases of COVID-19 suggests the possibility of liver damage, which could also impair amino acid and pyrimidine metabolisms since CPSI could also maintain pyrimidine pool. Notably, both GMP and carbamoyl phosphate show significant changes between fatal and mild patients, indicating that the disease progression is associated with immune dysfunction and nucleotide metabolism.

### Plasma lipidomic alternations correspond to clinical symptoms of COVID-19

We analyzed the lipids in different courses of fatal COVID-19 patients (F1-F4) that underwent significant change [F4 vs. H, >1 log_2_ FC <-1, typically *P* <0.05]. Most of the significantly changed lipids are up-regulated and a positive correlation between the alteration of lipids and the course of disease deterioration could be readily observed in the fatal patients (Figure 4A and Table S8). Lipid subclasses including diglycerides (DGs), free fatty acids (FAAs), and triglycerides (TGs), were identified in higher abundance in the fatality group (F vs. H), and the relative abundances of these lipids increased with the deterioration of the disease. Particularly, DG(16:0/20:2/0:0) exhibited the greatest log_2_ FC (+4.15) in DGs, and TG(14:0/22:1/22:3) exhibited the greatest log_2_ FC (+4.17) in all significantly altered TGs. The increases of DGs, FFAs, and TGs under pathological conditions have been previously reported. For instance, lipolysis of adipose tissue increases due to EBOV infection, which converts TG to FFA and DG, and also results in enhanced recycling of the fatty acids back into TGs (9).

**Figure 4.**
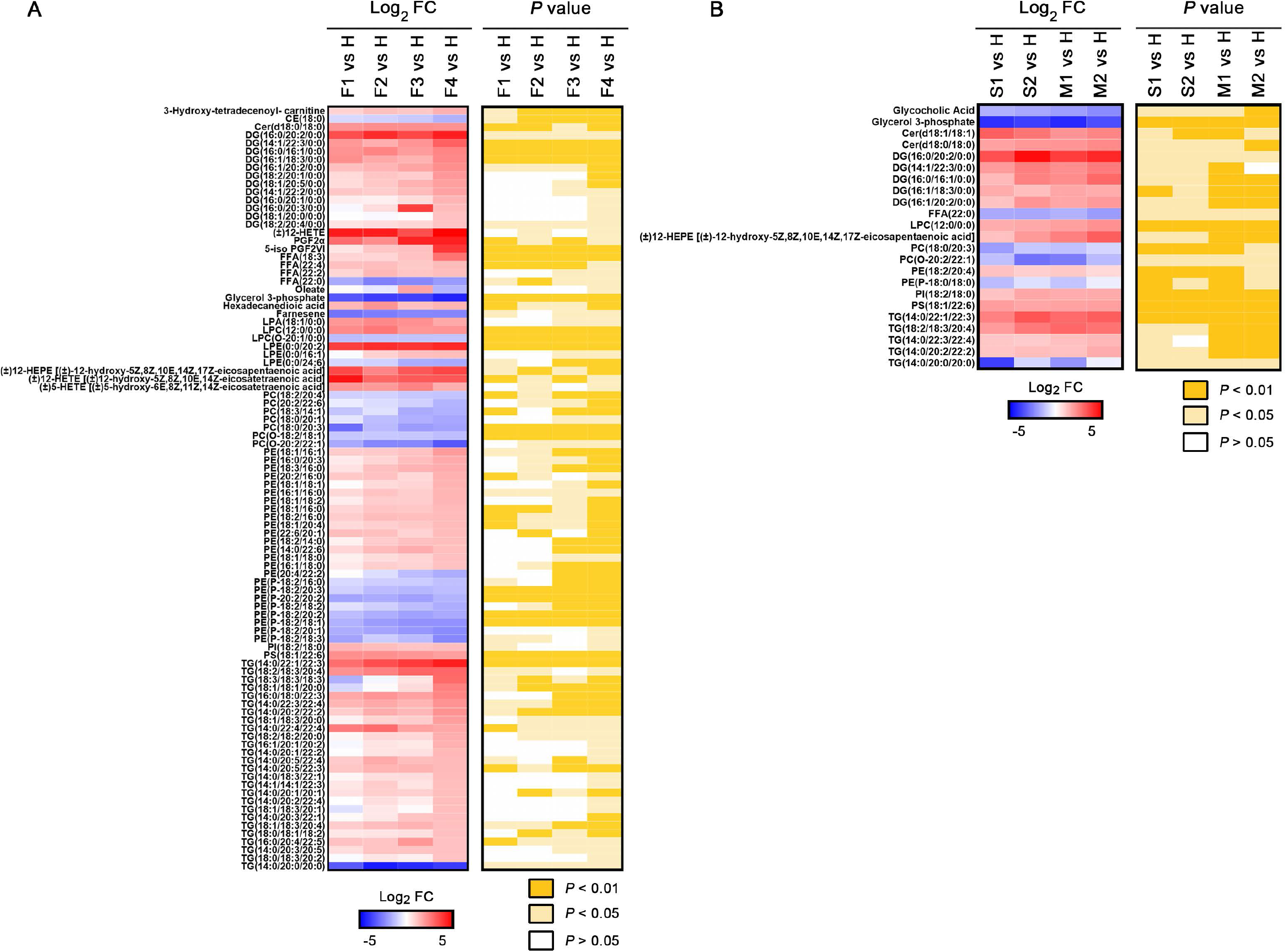
COVID-19 signatures in the plasma lipidome. Selected average plasma lipid expression levels and associated p values for COVID-19 fatality group vs. healthy volunteer group (A), and severe or mild vs. healthy groups (B). AA, arachidonic acid; BA, bile acid; CAR, carnitine; CE, cholesterol ester; Cer, ceramide; DG, diacylglycerides; TG, Triglycerides; FA, fatty acid; FFA, free fatty acids; LPA, lysophosphatidic acid; PC, phosphatidylcholine; LPC, lysophosphatidylcholine; PE, phosphatidylethanolamine; LPE, lysophosphatidyl ethanolamine; LPG, lysophosphatidylglycerol; PI, phosphatidylinositol; PS, phosphatidylserine; LPO, lipid peroxide.

Besides, we observed that phosphatidylcholines (PCs) were gradually reduced over the course of COVID-19 fatalities. PCs are synthesized in the liver and are the only phospholipid necessary for lipoprotein (18); therefore, the COVID-19-associated decrease of PCs in the fatality group indicates hepatic impairments happened in the fatality group. Additionally, decreases in lysophosphatidylcholines (LPCs) and PCs in blood plasma have been observed in sepsis, cancer, and Dengue infection (19-22).

We also analyzed the lipids in different courses of severe and mild COVID-19 patients (S1 and S2; M1 and M2) that underwent the significant change [S1 vs. H, >1 log_2_ FC <-1, typically *P* <0.05; M1 vs. H, >1 log_2_ FC <-1, typically *P* <0.05] (Figure 4B and Table S9). Similar to those of metabolites, the total numbers of significantly altered lipids (>1 log_2_ fold change (FC) <-1, typically *P* <0.05) in the severe and mild groups (S1 vs H, S2 vs H, M1 vs. H, and M2 vs H) were similar, which are significantly less than the number of altered lipids in the fatality group, indicating that the alterations of lipid metabolism were much more dramatic in fatal COVID-19 patients than in survivors. Besides, for either severe or mild groups of patients, many of their lipids had not returned to normal before their discharge from hospital (Figure 4B, S2 vs H and M2 vs H), even though SARS-CoV-2 could not be detected and the major clinical signs had disappeared in these patients based on official discharge criteria. Obviously, like the observations of metabolomic alterations, these discharged patients, no matter if they had experienced severe or mild symptoms, had not been fully recovered from the aftermath of COVID-19 in the aspects of both metabolite and lipid metabolisms.

Furthermore, the differentiating lipids were divided into those shared by all groups (F vs. H and S vs. H plus M vs. H) or those unique to the fatality group (F vs. H), and subsequently subjected to KEGG functional enrichment analysis. As shared by all the three symptomatic groups, differentiating lipids were enriched in total 7 pathways and significantly enriched in 4 pathways, including phosphatidylinositol signaling system, long-term depression, leishmaniasis, and inositol phosphate metabolism (Figure 5A-B and Table S10). In the case of fatality group, differentiating lipids were significantly enriched in 6 pathways, including retrograde endocannabinoid signaling, pathogenic *Escherichia coli* infection, Kaposi sarcoma-associated herpesvirus infection, glycosylphosphatidylinositol-anchor biosynthesis, glycerophospholipid metabolism and autophagy (Figure 5C-D and Table S11).

**Figure 5.**
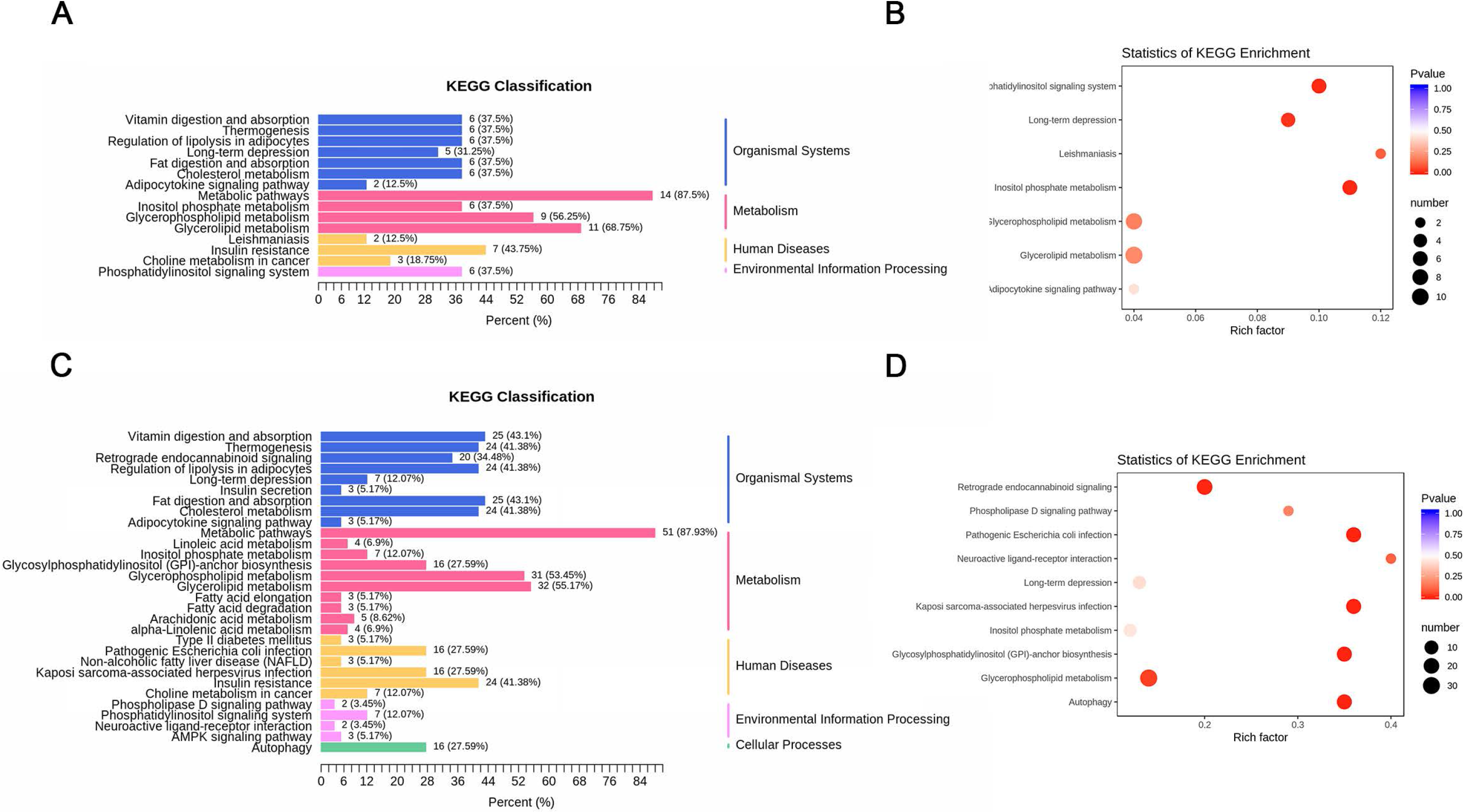
The lipidome KEGG enrichment analysis of COVID-19 patients. (A-B) KEGG pathway analysis of DEIs shared in all the groups. The color of bubbles represents the value of adjusted *P* value, and the size of bubbles represents the number of counts (sorted by gene ratio). (C-D) KEGG pathway analysis of DEIs shared unique to the fatal groups.

To highlight the top differentiating lipids, we showed 8 down-regulated lipids and 7 up-regulated lipids in COVID-19 patients compared with those in the healthy group (Figure 6 and Table S12), suggesting the dyslipidaemia in COVID-19 patients.

**Figure 6.**
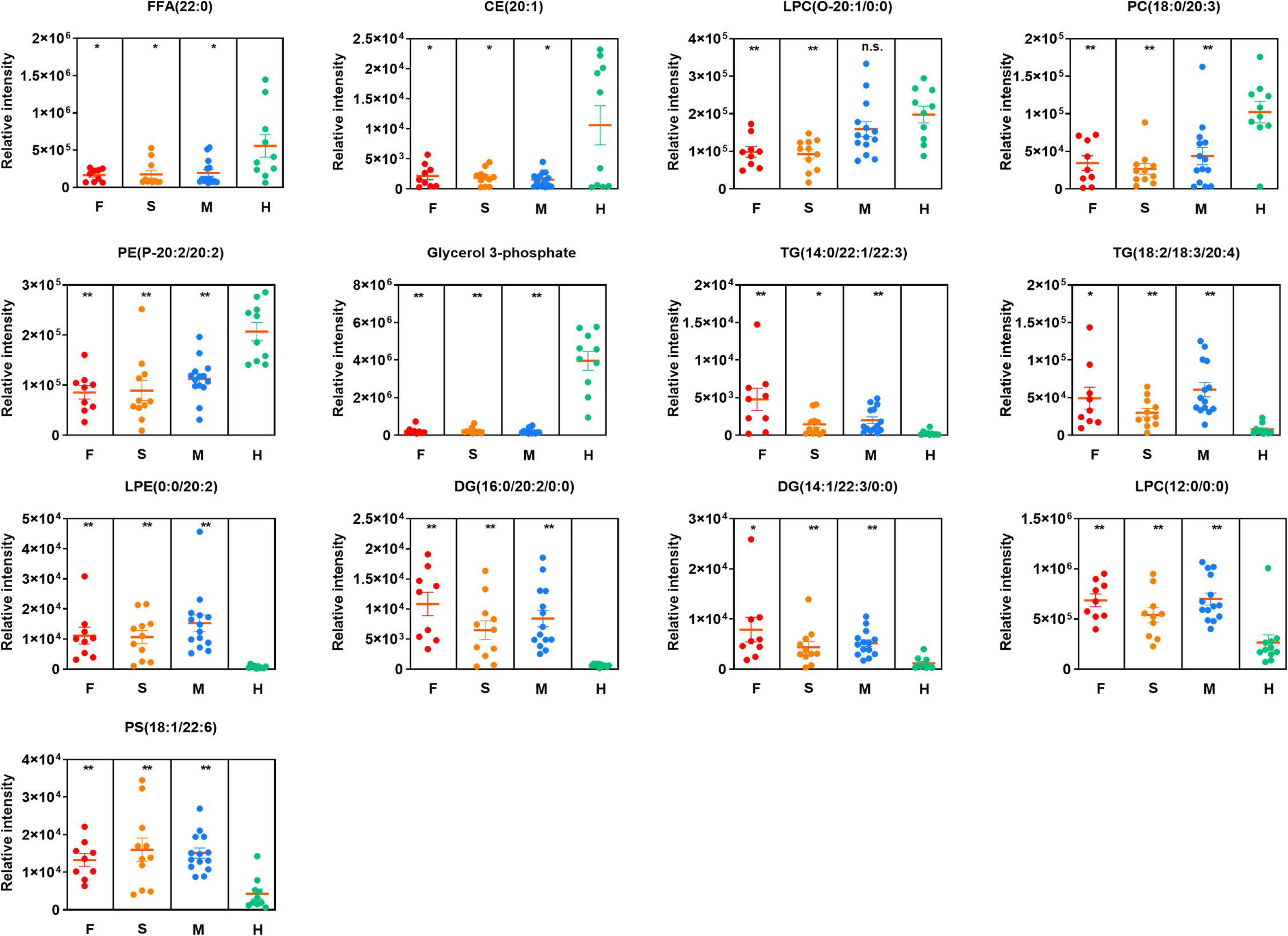
The potential lipidomic biomarkers of COVID-19. The relative intensity for each lipid. Each dot represents a patient sample, and each patient group is differently colored as indicated. F, fatalities; S, the patients with severe symptom; M, the patients with mild symptom; H, healthy volunteers. \**P*<0.05, \*\**P*<0.01.

### Logistic regression and receiver operating characteristic curve analysis

To rule out the possibility of potential biomarkers induced by age difference and/or gender disparity, we developed a logistic regression model for COVID-19 patients and healthy controls. As shown in Table 1, the *P*-value from either age or gender variable shows no statistically significant differences on metabolic alterations, even though patients were older with the progression of disease.

**Table 1.** Multivariable Analysis of the Associations of Age and Gender with COVID-19.

| **Variables** | **Odds Ratio** | **95% CI** | ***P*-value <sup>a</sup>** |
| --- | --- | --- | --- |
| **Age** | 0.95 | (0.89,1.01) | 0.14 |
| **Gender** | 3.67 | (0.80, 20.86) | 0.11 |

<sup>a</sup> *P* values were calculated using the 2-sided test.

Next, we generated ROC curves to assess the potential usefulness of plasma metabolite signatures for the diagnosis of COVID-19. Our ROC analyses revealed that combined five plasma metabolites were robust in discriminating COVID-19 patients from controls, with an area under the curve (AUC) value of 1.00 (data not shown). Then we analyzed ROC curve of each metabolite between COVID-19 and healthy subjects, revealing that malic acid and D-Xylulose 5-phosphate (Xu-5-P) show the best AUC values of 0.994 and 0.959, respectively (Figure 7A). Furthermore, the combined five plasma metabolite panel discriminating fatal group from mild group (Figure 7B), discriminating severe group from mild group (Figure 7C), and fatal group from severe group (Figure 7D) in ROC analysis, show the AUC values of 0.865, 0.708, and 0.737, respectively. Therefore, the combined five plasma metabolites could be a useful panel for COVID-19 diagnosis. Moreover, we further generated ROC curves of down-regulated lipids (Figure 7E) and up-regulated lipids (Figure 7F) for discriminating COVID-19 patients and healthy controls, respectively. Intriguingly, ROC curve analysis of glycerol 3-phosphate with an AUC value of 1.00 was observed, suggesting that circulating glycerol 3-phosphate would be a good biomarker for COVID-19. These obtained results suggest metabolomics and lipidomics provide a potential tool for disease diagnosis and drug targets in the current pandemic.

**Figure 7.**
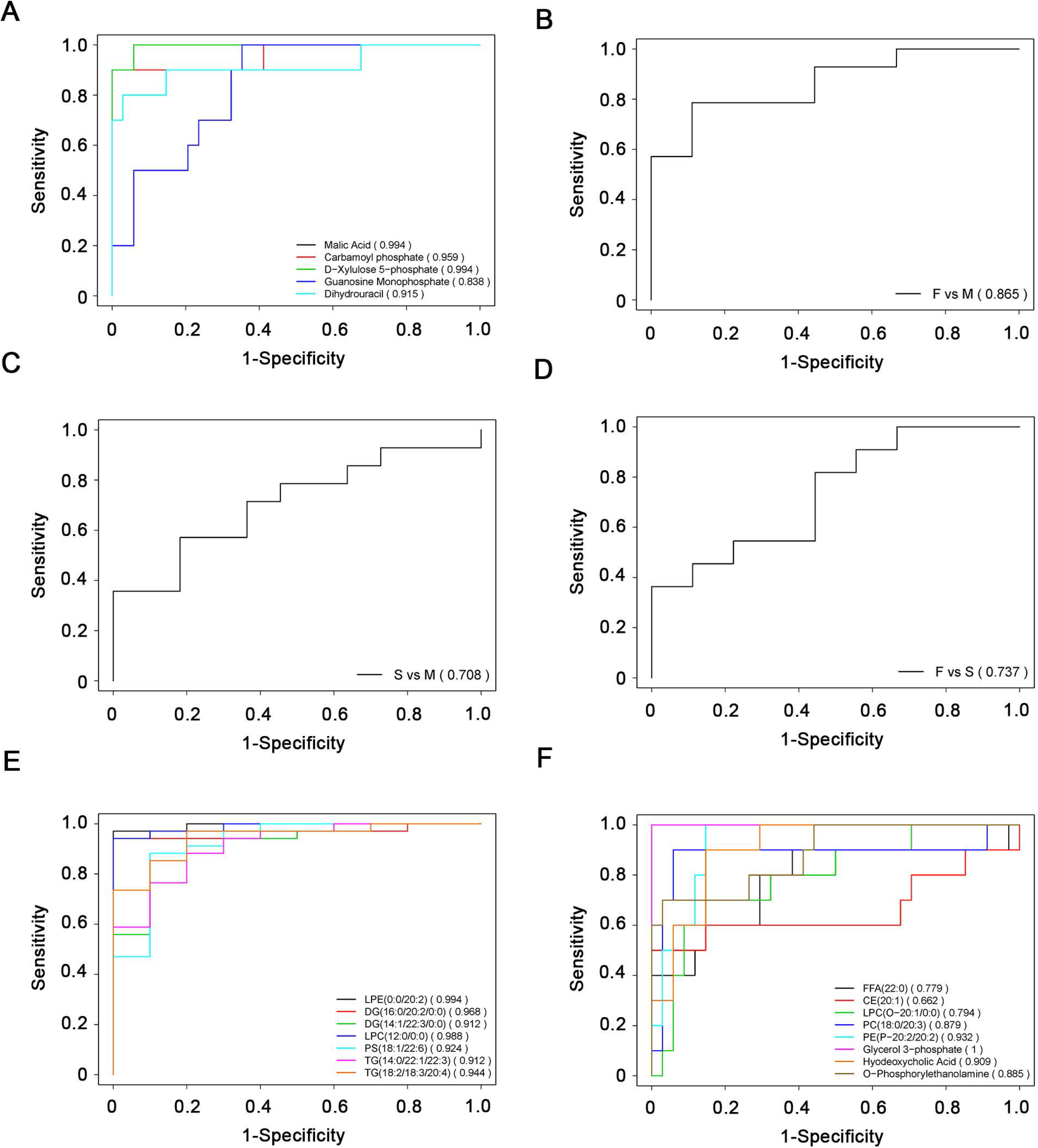
ROC curve analysis for the predictive power of biomarkers for distinguishing COVID-19 patients and healthy controls. (A) ROC curve analysis for the predictive power of each plasma metabolite for distinguishing COVID-19 groups from healthy controls; (B) ROC curve analysis for the predictive power of combined five plasma metabolites for distinguishing fatal group from mild group; (C) ROC curve analysis for the predictive power of combined five plasma metabolites for distinguishing severe group from mild group; (D) ROC curve analysis for the predictive power of combined five plasma metabolites for distinguishing fatal group from severe group; (E) ROC curve analysis for the predictive power of combined down-regulated lipids for distinguishing fatal group from healthy controls; (F) ROC curve analysis for the predictive power of combined up-regulated lipids for distinguishing fatal group from healthy controls.

## Discussion

The main purposes of this study were to generate a high-quality resource of metabolomic and lipidomic datasets associated with COVID-19 and identify the potential biomarkers for disease diagnosis for a better understanding of the pathogenesis of COVID-19.

Among the highlighted biomarkers, malic acid and glycerol 3-phosphate showed the greatest reduction when comparing the fatality patients with healthy volunteers, and also showed dramatic reduction in both severe and mild groups. Malic acid has important physiological functions, as it can directly enter the circulation of TCA cycle to participate in human energy metabolism. Besides, malic acid can accelerate ammonia transformation to lower ammonia concentration in liver and to protect liver (23, 24). Therefore, the dramatic reduction of malic acid is consistent with the hepatic impairment associated with COVID-19. Moreover, malic acid has been found to protect endothelial cells of human blood vessels and resist damage to endothelial cells.

Xu-5-P is a metabolite of the pentose phosphate pathway that mediates the effects of carbohydrate feeding on the glycolytic pathway, as well as fatty acid and triglyceride synthesis. Xu-5-P is the coordinating signal that both activates phosphofructokinase in glycolysis and promotes transcription of the genes for lipogenesis, the hexose monophosphate shunt, and glycolysis, and is required for *de novo* synthesis of fat and hepatic energy utilization (25-28). The reduction of Xu-5-P indicates that altered glucose and lipid metabolisms are also a reflection of hepatic impairment.

Carbamoyl phosphate is an important intermediate metabolite involved in removing excess ammonia in the urea cycle (14, 15). This metabolite is the downstream product of CPSI in mitochondria of liver cells. The observed down-regulation of carbamoyl phosphate levels is associated with the severity of COVID-19, as its level in the mild patients were affected in the least extent. Importantly, the dramatic reduction of carbamoyl phosphate is usually associated with urea cycle disorder, raising the concern about the possibility of hyperammonemia and liver failure in COVID-19 patients. The postulation seems to be consistent with deficiency of malic acid as mentioned above. In addition, the metabolisms of purine and thyroid hormones were significantly altered in the fatality group. Purine metabolism mainly occurs in human liver, and the thyroid hormone can affect hepatic protein synthesis and glycogen decomposition. Therefore, our findings show that the development of COVID-19 can cause hepatic impairment in these patients, which is consistent with the observations that a large number of COVID-19 patients showed liver function abnormalities (Table S13) (6).

Besides, the reductions of dihydrouracil, an intermediate breakdown product of uracil and guanosine monophosphate (GMP) (29), are proposed to be caused by the defects of human metabolism. It should be noted that GMP production is not only mediated by GMP synthase but also CD39 and CD73. Indeed, CD39/CD73 axis plays a crucial role in immunity and inflammation (30). Another metabolite glycerol 3-phosphate is a conserved three-carbon sugar and an obligatory component of energy-producing reactions including glycolysis and glycerolipid biosynthesis (31). Moreover, glycerol 3-phosphate is an important mobile regulator of systemic acquired resistance, which provides broad spectrum systemic immunity in response to pathogenic infections (32). These metabolites show good correlation with the progress and severity of COVID-19, and can therefore serve as biochemical indicators for immune dysfunction of this disease.

Meanwhile, the lipids involved in glycerol metabolism pathway were upregulated, which maintain the balance of the energy metabolites of the body and are beneficial to the energy required for viral replication, suggesting that SARS-CoV-2 probably hijacks cellular metabolism like many other viruses (33). Our data show that the metabolic pathway of glycerophospholipids has been significantly changed, and glycerophospholipids are closely related with cardiovascular diseases. However, we did not find obvious pattern or significant difference of underlying diseases, such as hypertension, cardiac disease, diabetes, cerebrovascular disease, chronic hepatitis, and cancer, in the medical records of all the patient groups involved in this study. Therefore, this finding suggests that the fatality caused by COVID-19 might be related with cardiac impairment. Interestingly, COVID-19 has been recently reported to probably cause the loss of the smell and taste sense (https://www.npr.org/sections/goatsandsoda/2020/03/26/821582951/is-loss-of-smell-and-taste-a-symptom-of-covid-19-doctors-want-to-find-out), and the KEGG analysis also showed that the taste transduction pathway is affected.

The metabolomic and lipidomic analyses also show that, although the patients in both the severe and mild symptom groups had met the official hospital discharge criteria as their COVID-19 nucleic acid tests for turning out to be negative consecutively twice and major clinical signs disappeared, many of their fundamental metabolites and lipids failed to return to normal. This finding suggests that these discharged patients, regardless of the severity of their previous symptoms, had not been fully recovered from the disease in the aspect of metabolism, particularly hepatic functions. Therefore, even after the clearance of SARS-CoV-2 from patient bodies, these convalescent COVID-19 patients still need better nutrition and care that would be very helpful for their faster and full recovery from the disease.

The metabolomic and lipidomic alterations in patient plasma mainly reflect the systematic responses of the metabolisms of diverse cell types and organ systems that were affected by SARS-CoV-2. Therefore, the interpretations of the datasets should be integrated with other types of system studies, such as the transcriptome and proteome of specific tissue and body fluid samples, as well as clinical observations and laboratory examinations, to have a clearer and more comprehensive picture of the development of this disease. Moreover, such an integration would help us to better understand the impacts of COVID-19 to specific cells and/or tissues infected by SARS-CoV-2.

In summary, the metabolomic and lipidomic datasets of the cohort of COVID-19 patients under different symptomatic conditions are highly valuable resources for a better understanding of the host metabolic responses associated with COVID-19, which expands our knowledge about the pathogenesis of COVID-19, accelerates identification of disease biomarkers and development of diagnostic assays, and provides hints of potential therapeutic strategies.

## Materials and Methods

Materials and methods are detailed described in Supplementary data.

## Data Availability

All data are available from the authors upon request

## Supplementary data

Supplementary methods and materials

Supplementary Figure S1-S4

Supplementary Table S1-S15

## Author contributions

D.W., T.S., X.Y. and J.-X.S. performed experiments with the help of W.L., M.H., Y.Y., Q.Y., T.Z., J.X., Y.W., J.M., H.W., T.T., Y.R. Y.W.; M.Z., C.Y. and S.-H.L. analyzed the metabolomics and lipidomics data with the help of D.W. and Y.Q.; S.-H.L., Y.Q., D.-Y.Z., Y.S., and X.Z. performed the experimental design and data interpretation; X.Z, Y.Q., Y.S., D.-Y.Z. and S.-H.L. analyzed the data and wrote the paper; X.Z, Y.Q., Y.S. and D.-Y.Z. designed and supervised the overall study.

## Acknowledgments

We thank the patients, and the nurses and clinical staffs who are providing care for these patients. We thank the helpful discussions with Drs. Yan Wang and Yong Liu at Wuhan University. We also thank many staff members at Wuhan Jinyintan Hospital and Wuhan Metware Biotechnology Co., Ltd. for their contributions and assistance in this study. We sincerely pay tribute to our colleagues who have strived in the forefront of taking care of COVID-19 patients and are studying this novel coronavirus in Wuhan and other places around the world.

This work was supported by the Strategic Priority Research Program of CAS (XDB29010300 to X.Z.), the National Science and Technology Major Project (2020ZX09201-001 to D.-Y.Z, and 2018ZX10101004 to X.Z.), National Natural Science Foundation of China (81873964 to Y.Q., 31670161 to X.Z., 81971818, and 81772047 to Y.S.), the Fundamental Research Funds for the Central Universities (20720200013 to S-H.L.), and Grant from Clinical Research Center for Anesthesiology of Hubei Province (No.2019ACA167).

## Supplementary Data

**Supplementary methods and materials**

**Supplementary Figure S1-S4**

**Supplementary Table S1-S15**

## Methods and Materials

### Ethics and Human Subjects

All work performed in this study was approved by the Wuhan Jinyintan Hospital Ethics Committee and written informed consent was obtained from patients. Diagnosis of SARS-CoV-2 infection was based on the New Coronavirus Pneumonia Prevention and Control Program (6th edition) published by the National Health Commission of China. Healthy subjects were recruited from healthcare workers and laboratory workers at Wuhan Jinyintan Hospital and Wuhan Institute of Virology, CAS, none of whom had previously experienced SARS-CoV-2 infection.

### Patient Samples

SARS-CoV-2-positive patients were enrolled in the study after diagnosis. Blood sample (≤3mL) from fatal COVID-19 patients were collected over the course of their disease at intervals of 3-5 days. Blood sample (≤3mL) from the patients with severe and mild symptoms were collected at the time when the disease were most serious (3-7 days after hospitalization) and the time before discharge. Single samples were collected from healthy volunteers recruited from healthcare workers and laboratory workers at Wuhan Jinyintan Hospital and Wuhan Institute of Virology. The throat swabs and serological testing of healthy volunteers were negative for SARS-CoV-2. All blood samples were collected after fasting overnight and by potassium-EDTA blood collection tubes. All samples used in this study are described in Table S1. All the blood samples were treated according to the biocontainment procedures of the processing of SARS-CoV-2-positive sample.

### Methods for extraction of hydrophilic and hydrophobic compounds

To analysis hydrophilic compounds, sample was thawed on ice, 3 volumes of ice-cold methanol was added to 1 volume of plasma/serum, whirled the mixture for 3 min and centrifuge it with 12,000 g at 4°C for 10 min. Then the supernatant was centrifuged at 12,000 g at 4°C for 5 min, and then collected the supernatant and subjected them to LC-MS/MS analysis.

To analysis hydrophobic compounds, sample was thawed on ice, whirl around 10 s, and then centrifuge it with 3000 g at 4°C for 5 min. Take 50 μL of one sample and homogenized it with 1mL mixture (include methanol,MTBE and internal standard mixture). Whirled the mixture for 2 min. Then added 500 μL of water and whirled the mixture for 1 min, and centrifuged it with 12,000 g at 4°C for 10 min. Extracted 500 μL supernatant and concentrated it. Dissolved powder with 100 μL mobile phase B and subjected to LC-MS/MS analysis.

### UPLC conditions of hydrophilic and hydrophobic compounds

The sample extracts of hydrophilic compounds were analyzed using an LC-ESI-MS/MS system (UPLC, Shim-pack UFLC SHIMADZU CBM A system, MS, QTRAP® 6500+ System). The samples were injected onto a Waters HSS T3 column (1.8 µm, 2.1 mm×100 mm). Column temperature, flow rate and injection volume were set 40°C, 0.4 mL/min and 2μL, respectively. Mobile phase was composed of water containing 0.1% formic acid (A) and acetonitrile containing 0.1% formic acid (B). The gradient program initiated from 5% B increased to 90% B in 11.0 min, and held for 1 min and then decreased 5% B for re-equilibrium.

The sample extracts of hydrophobic compounds were analyzed using an LC-ESI-MS/MS system (UPLC, Shim-pack UFLC SHIMADZU CBM A system, MS, QTRAP® 6500+ System). The samples were injected onto a Thermo C30 column (2.6 μm, 2.1 mm×100 mm). Mobile phase was composed of acetonitrile/water (60/40, v/v) containing 0.04% acetic acid and 5 mmol/L ammonium formate (A) and acetonitrile/isopropanol (10/90, v/v) containing 0.04% acetic acid and 5 mmol/L ammonium formate (B). The gradient program initiated from 20%B to 50% in 3 min, to 65% in 2 min, to 75% 4 min and to 90% in 6.5 min. The flow rate, column temperature and injection volume were set 0.35 ml/min, 45°C and 2μL, respectively. The effluent was alternatively connected to an ESI-triple quadrupole-linear ion trap (QTRAP)-MS.

### ESI-Q TRAP-MS/MS of hydrophilic and hydrophobic compounds

LIT and triple quadrupole (QQQ) scans were acquired on a triple quadrupole-linear ion trap mass spectrometer (QTRAP), QTRAP® LC-MS/MS System, equipped with an ESI Turbo Ion-Spray interface, operating in positive and negative ion modes and controlled by Analyst 1.6.3 software (Sciex). The ESI source operation parameters were as follows: ion source, turbo spray; source temperature 550 °C; ion spray voltage (IS) 5500 V in positive ion mode (or −4500 V in negative ion mode); ion source gas I (GSI), gas II (GSII), curtain gas (CUR) were set at 45, 55, and 35 psi, respectively; the collision gas (CAD) was medium. Instrument tuning and mass calibration were performed with 10 and 100 μmol/L polypropylene glycol solutions in QQQ and LIT modes, respectively. QQQ scans were acquired as MRM experiments with collision gas (nitrogen) set to 5 psi. Declustering potential (DP) and collision energy (CE) for individual MRM transitions was done with further DP and CE optimization. A specific set of MRM transitions were monitored for each period according to the metabolites within this period. Each sample analysis was conducted by both positive and negative ion modes, and the MRM transitions were listed in Table S15.

### Plasma mentalities and lipids data analysis

The mass spectrum data were processed by Software Analyst 1.6.3. The repeatability of metabolite extraction and detection can be judged by total ion current (TIC) and multi peak MRM. Based on home-made MWDB (metadata database) and other databases, qualitative analysis of information and secondary general data was carried out according to retention time (RT) and mass-to-charge ratio. Metabolite structure analysis referred to some existing mass spectrometry public databases, mainly including massbank (http://www.massbank.jp/), knapsack (http://kanaya.naist.jp/knapsack/), HMDB (http://www.hmdb.ca/), and Metlin (http://metlin.scripps.edu/index.php). The metabolite identification was conduced by reference standards in our home-made database and public databases, and more detailed information is listed in Table S14.

For the quality control (QC) of metabolomic analysis, we pipette 10 μL of each sample to pool a QC sample. When running sample sets on column, one QC sample was injected after 10 samples in the sequence. Metabolite quantification was accomplished by using multiple reaction monitoring (MRM) of triple quadrupole mass spectrometry. Opened the mass spectrum file under the sample machine with multiquant software to integrated and calibrated the chromatographic peaks. The peak area of each chromatographic peak represented the relative content of the corresponding substance. Finally, exported all the integral data of chromatographic peak area to save, and used the self-built software package to remove the positive and negative ions of metabolites. We calculated coefficient of variation (CV) values of the metabolites in QC samples, and removed the metabolites whose CV values were larger than 0.5. When the metabolites were detected in both positive and negative ionization modes, we removed the metabolites with larger CVs in either positive or negative mode.

To maximize identification of differences in metabolic profiles between groups, the orthogonal projection to latent structure discriminant analysis (OPLS-DA) model was applied using the MetaboAnalyst online tool (https://www.metaboanalyst.ca/). The OPLS-DA model was evaluated with the relevant R^2^ and Q^2^. And we used the permutation to assess the risk that the current OPLS-DA model is spurious.

### Pathway Enrichment

We used the Kyoto Encyclopedia of Genes and Genomes (KEGG) database (http://www.genome.jp/kegg/) to analyze the KEGG pathway enrichment to identify highly enriched metabolic pathways in differential metabolites or lipids. The p-value <0.05 was considered significantly changing pathways and was used for subsequent analysis.

### Statistically Processed Datasets

Plasma metabolomics and lipidomics datasets (including fold-change and *P*-values for various group comparisons) are provided in Table S4-S5 and S8-S9. Plasma metabolomics pathway enrichment are provided in Table S6-S7. Plasma lipidomic pathway enrichment are provided in Table S10-S11.

### Raw Data

All raw LC-MS/MS data has been deposited to the iProX under the accession number: PXD018307.

### Statistics

The orthogonal-projection-to-latent-structure–discriminant-analysis (OPLS-DA) model was applied using R package “MetaboAnalyst”. And the OPLS-DA model verification was performed by a permutation test repeated 200 times. In general, P < 0.05 indicated the available OPLS-DA model. Student *t* test and fold change were also applied to measure the significance of each metabolite. Statistical significance was analyzed using one-tailed Student’s *t* test or Fisher’s exact test, and *P* < 0.05 was considered to be statistically significant. The *P* value was corrected for multiple testing via false-discovery rate (FDR) using the Benjamini-Hochberg method.

Logistic regression analysis and receiver operating characteristic (ROC) analysis were used for diagnosis of COVID-19 patient samples and healthy subjects. ROC curves were utilized to evaluate the biomarkers performance. It was conducted applying R software version 3.6.1. (R Foundation for Statistical Computing, Vienna, Austria).

## Supplementary Figure legends

**Figure S1.**
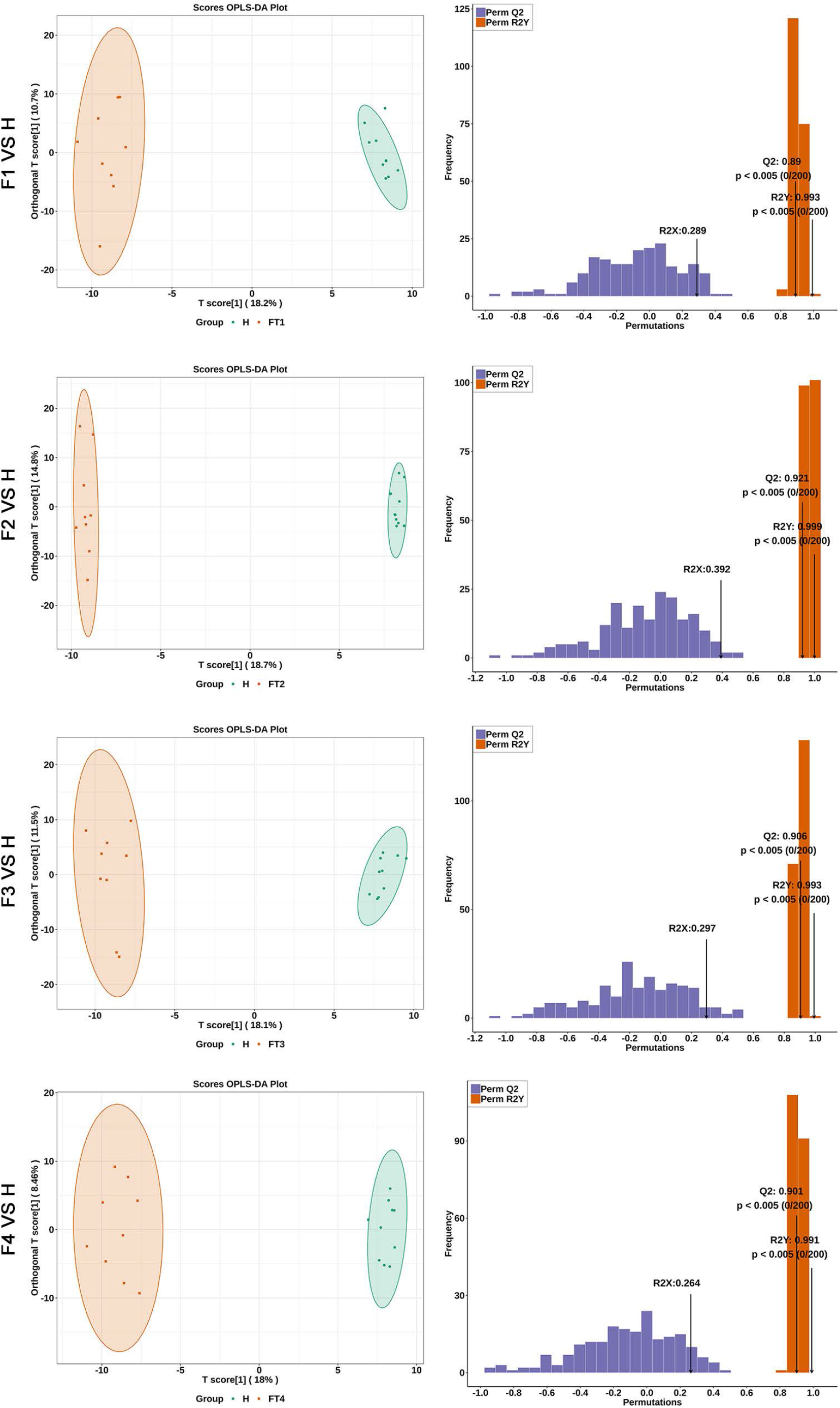
The orthogonal projection to latent structure discriminant analysis (OPLS-DA) showed the best possible discrimination of metabolites between fatal COVID-19 patients and healthy people as indicated. The x-axis represents the prediction component that shows differences between groups, and the y-axis represents the orthogonal component differences within the group. R2 represents goodness of fit, Q2 represents goodness of prediction, and *P* value shows the significance level of the model (x-axis = predictive components, y-axis = orthogonal component).

**Figure S2.**
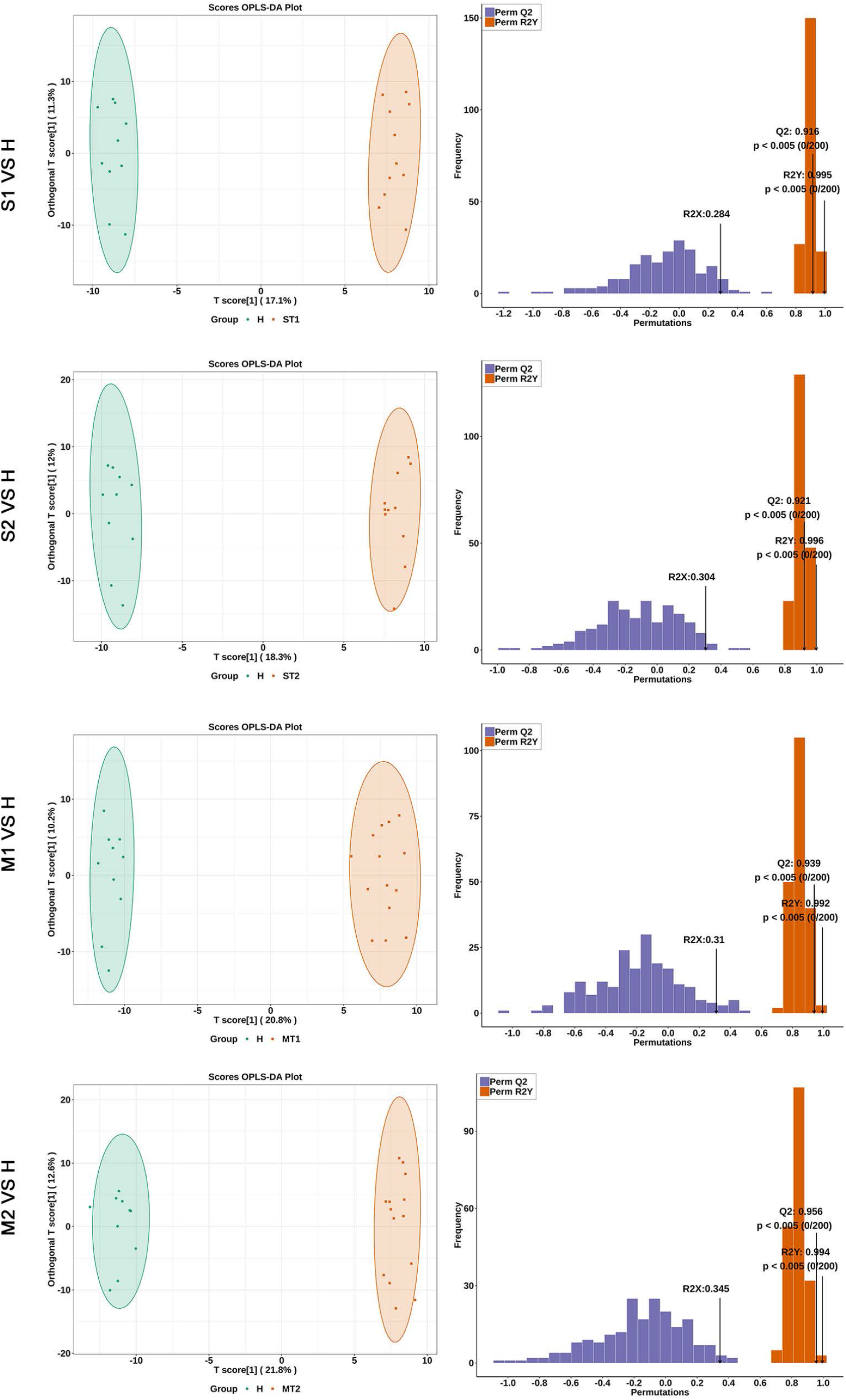
The OPLS-DA showed the best possible discrimination of metabolites between severe or mild COVID-19 patients and healthy people as indicated. The x-axis represents the prediction component that shows differences between groups, and the y-axis represents the orthogonal component differences within the group. R2 represents goodness of fit, Q2 represents goodness of prediction, and *P* value shows the significance level of the model (x-axis = predictive components, y-axis = orthogonal component).

**Figure S3.**
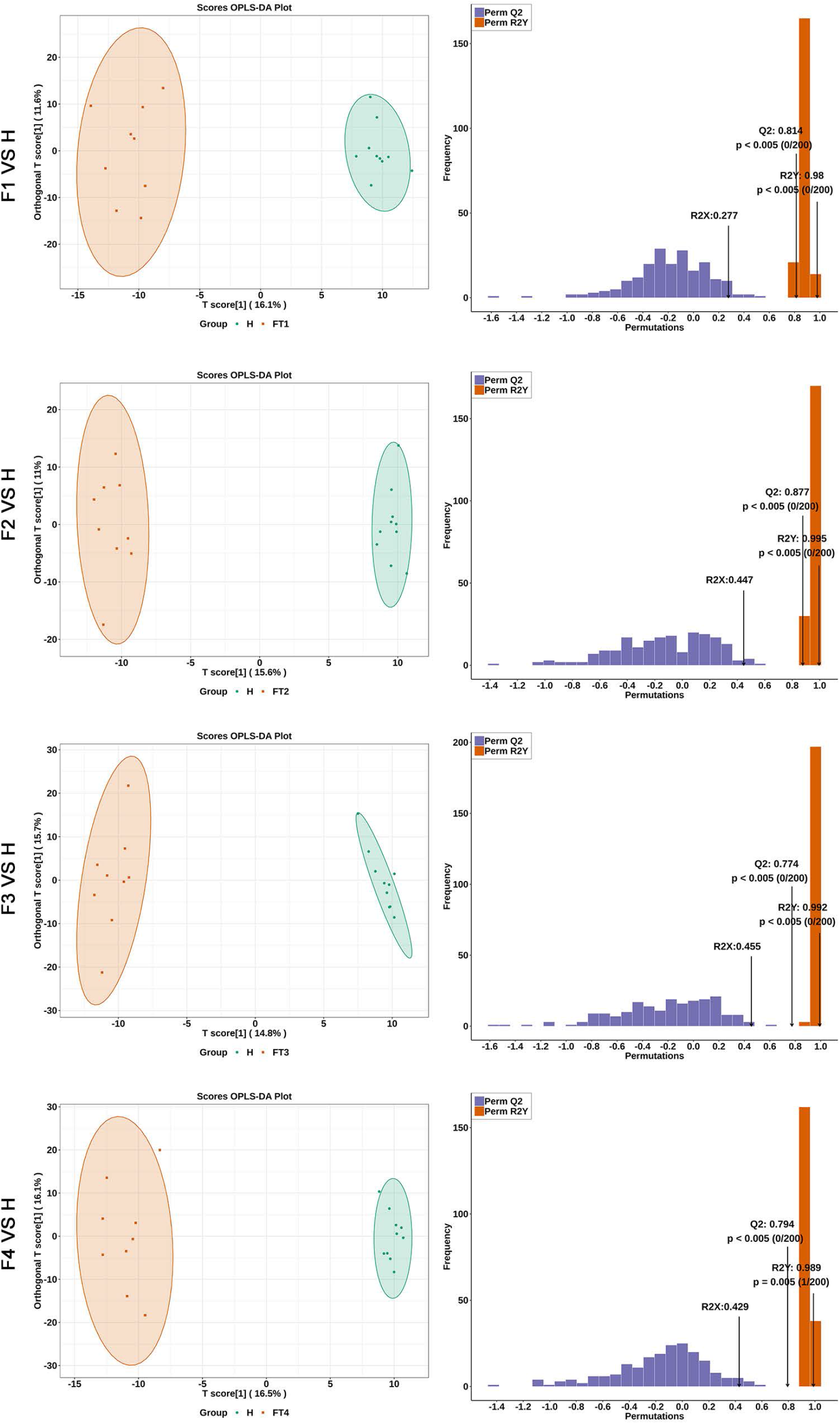
The OPLS-DA showed the best possible discrimination of lipids between fatal COVID-19 patients and healthy people as indicated. The x-axis represents the prediction component that shows differences between groups, and the y-axis represents the orthogonal component differences within the group. R2 represents goodness of fit, Q2 represents goodness of prediction, and *P* value shows the significance level of the model (x-axis = predictive components, y-axis = orthogonal component).

**Figure S4.**
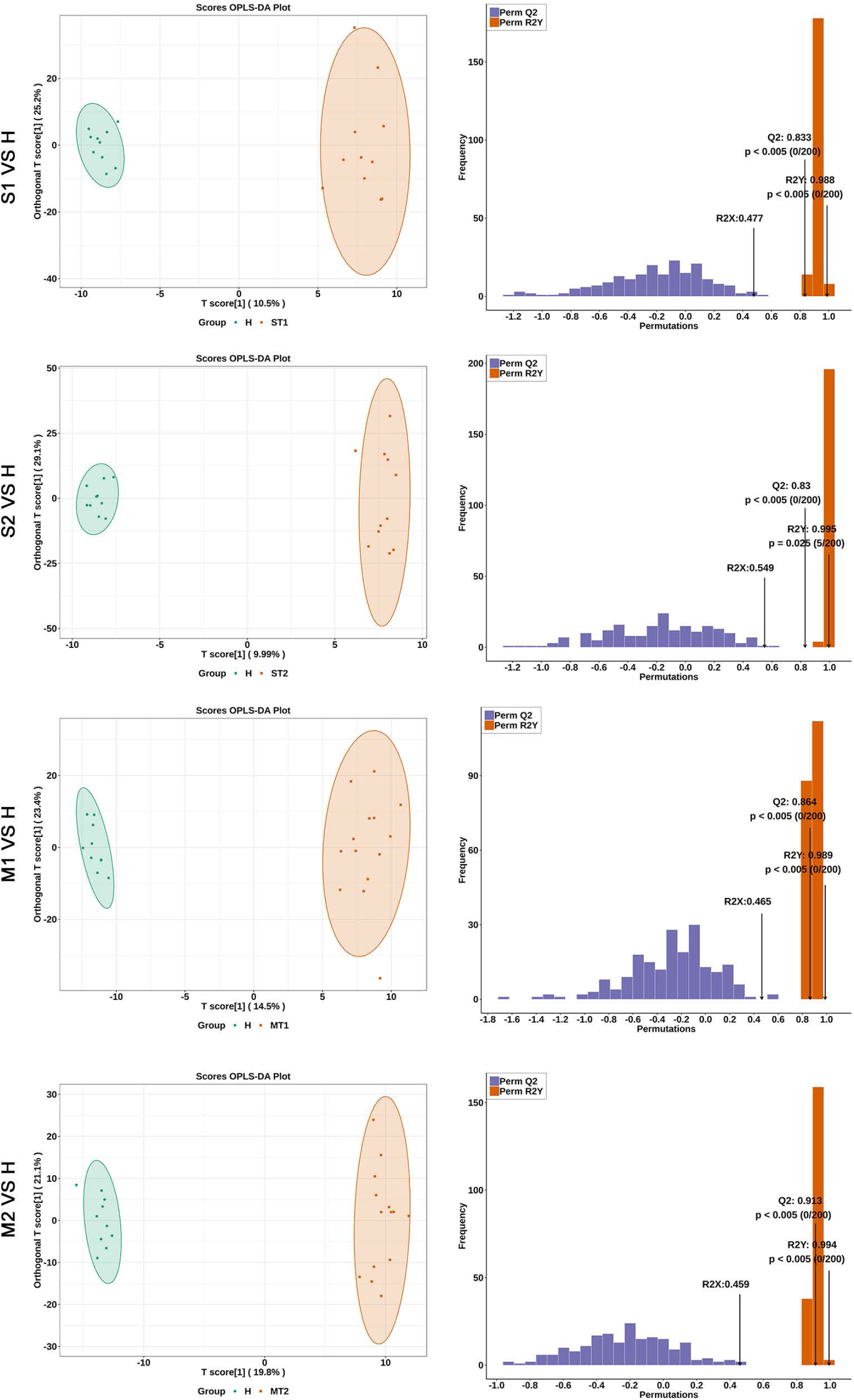
The OPLS-DA showed the best possible discrimination of lipids between severe or mild COVID-19 patients and healthy people as indicated. The x-axis represents the prediction component that shows differences between groups, and the y-axis represents the orthogonal component differences within the group. R2 represents goodness of fit, Q2 represents goodness of prediction, and *P* value shows the significance level of the model (x-axis = predictive components, y-axis = orthogonal component).

**Table S1.**
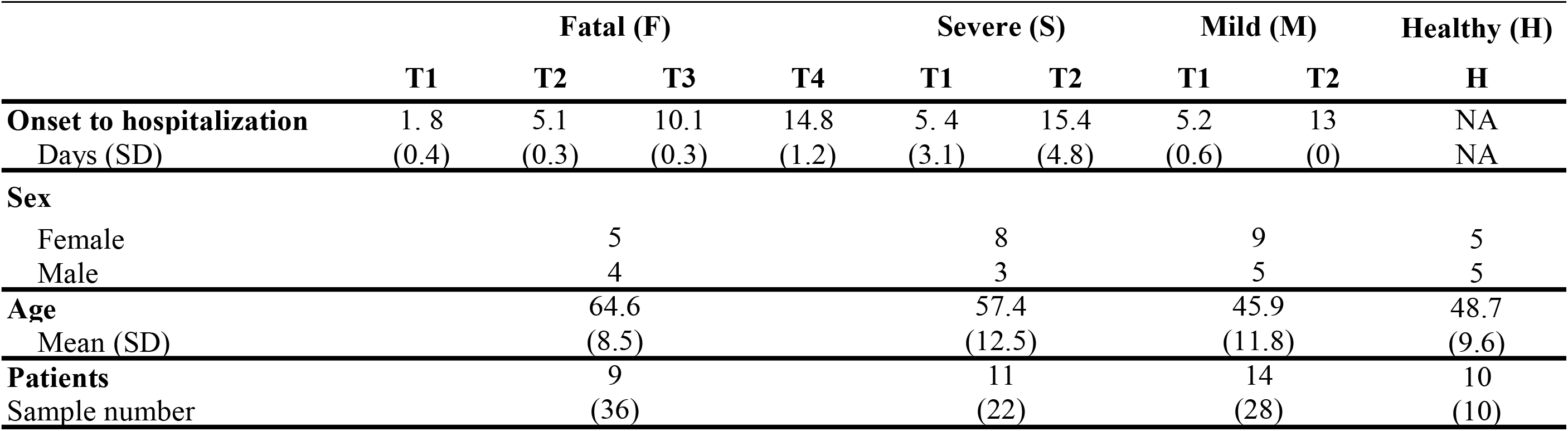
Study design and patients.

**Table S2.**
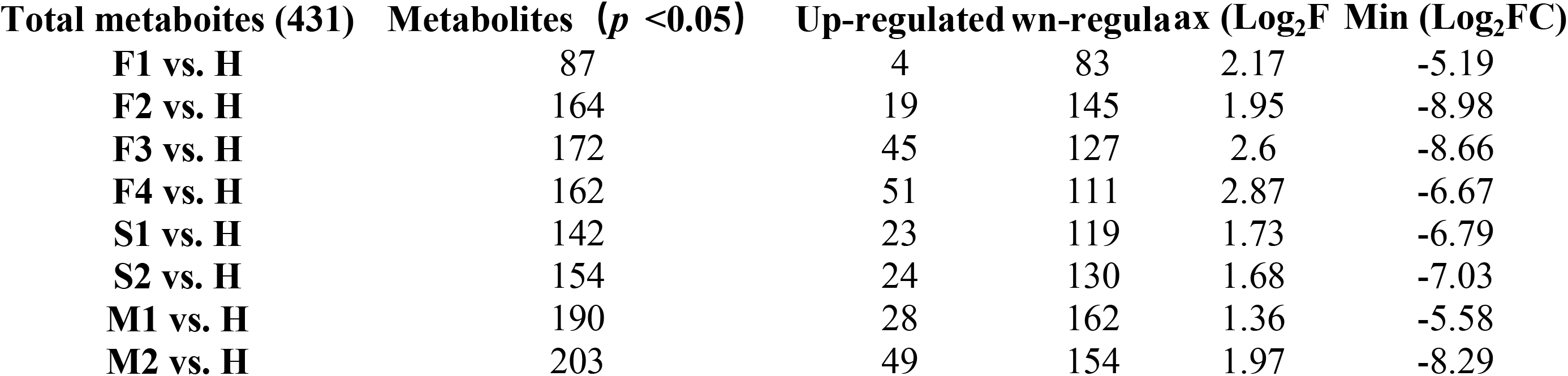
Overview of total changed metaboites.

**Table S2.** Overview of total changed lipids.

| **Total lipid (698)** | **Lipids ($p < 0.05$)** | **Up-regulated** | **Down-regulated** | **Max (Log<sub>2</sub>FC)** | **Min (Log<sub>2</sub>FC)** |
| --- | --- | --- | --- | --- | --- |
| **F1 vs. H** | 255 | 111 | 144 | 4.54 | -3.02 |
| **F2 vs. H** | 203 | 134 | 69 | 4.2 | -3.6 |
| **F3 vs. H** | 221 | 135 | 86 | 4.23 | -3.6 |
| **F4 vs. H** | 248 | 152 | 96 | 2.29 | -4.1 |
| **S1 vs. H** | 157 | 57 | 100 | 5.37 | -4.08 |
| **S2 vs. H** | 158 | 104 | 54 | 4.76 | -3.78 |
| **M1 vs. H** | 120 | 82 | 38 | 4.44 | -4.23 |
| **M2 vs. H** | 127 | 93 | 34 | 5.61 | -3.47 |

**Table S4.**
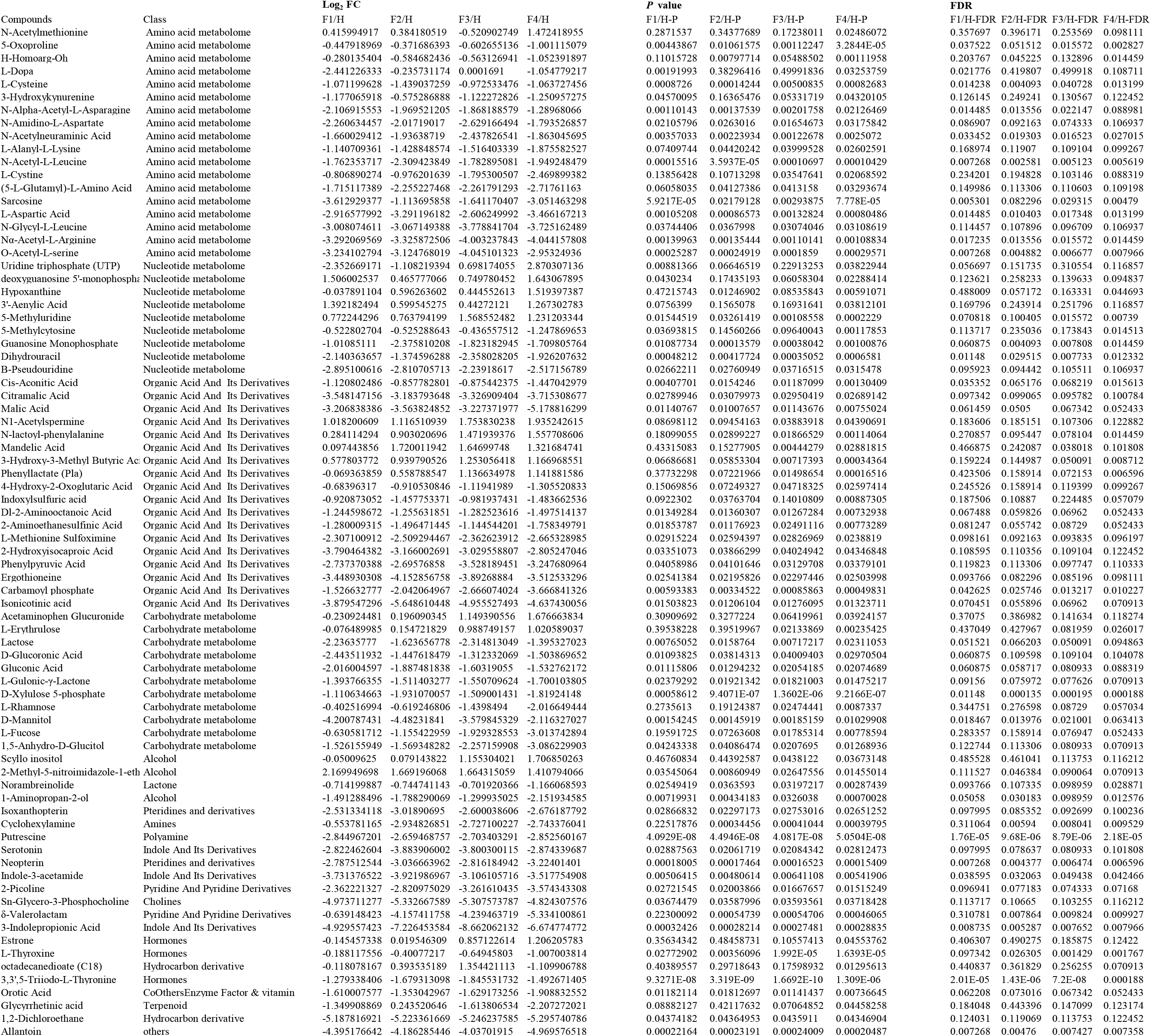
Metabolomics data of F vs H.

**Table S5.**
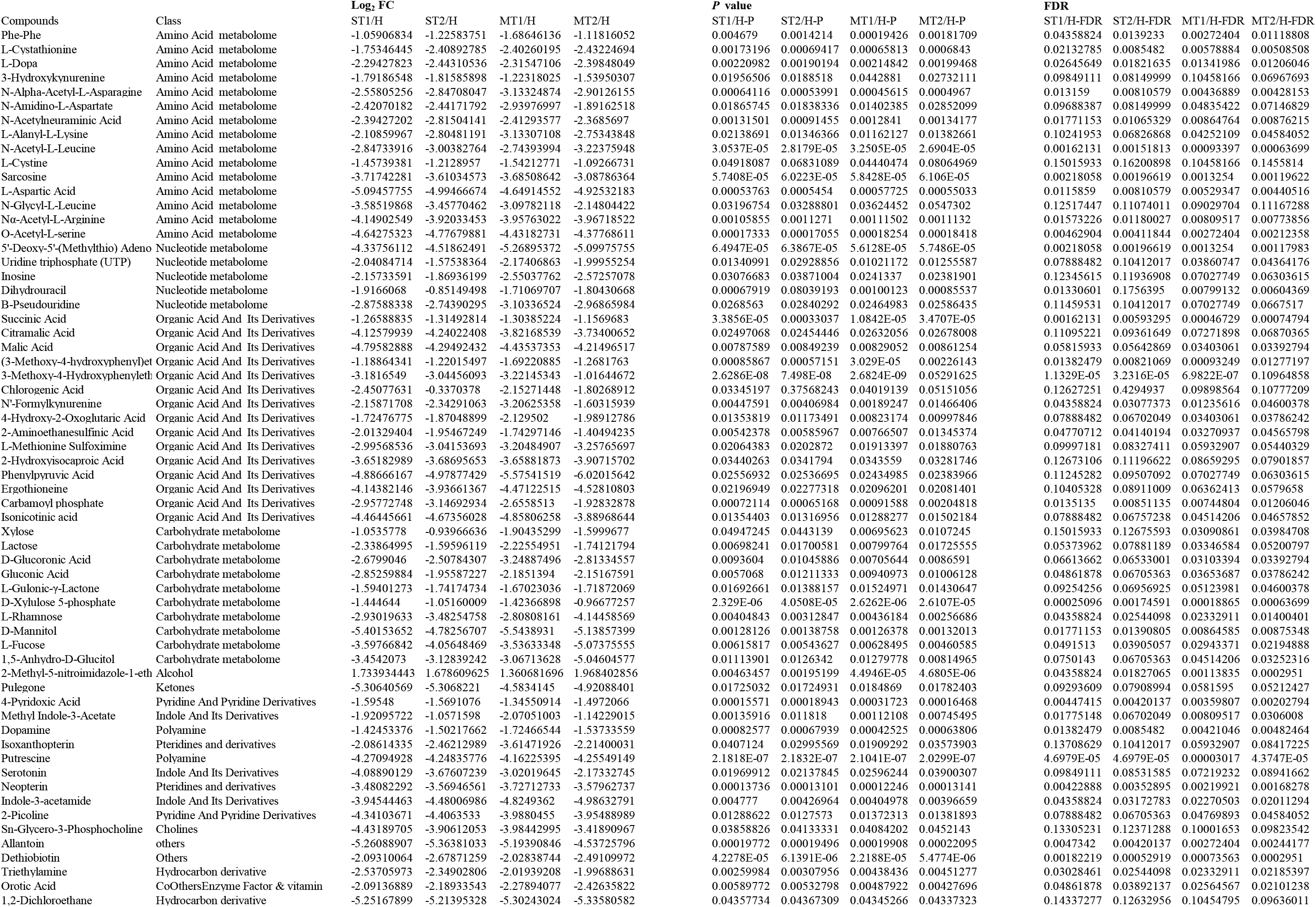
Metabolomics data of S vs H and M vs H.

**Table S6.**
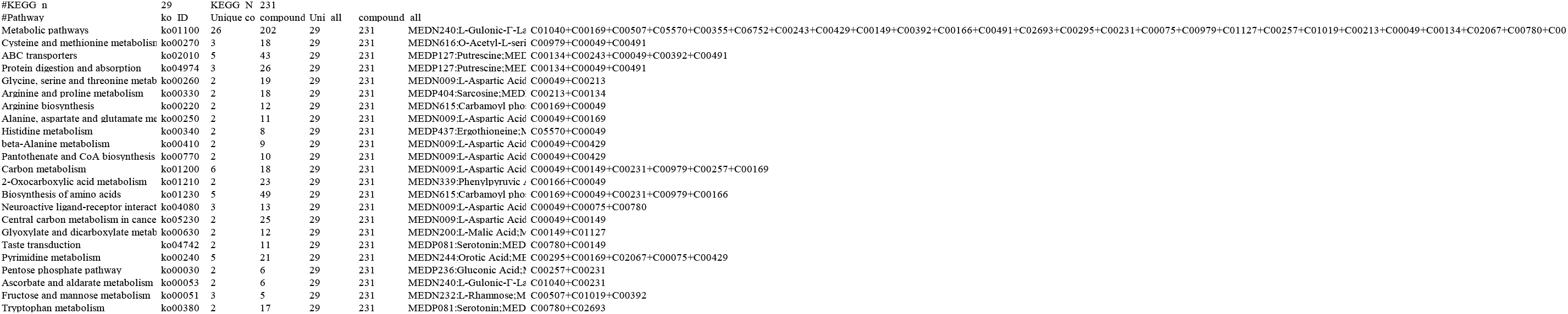
KEGG enrichment analysis of DEMs shared by F vs H, S vs H and M vs H.

**Table S7.**
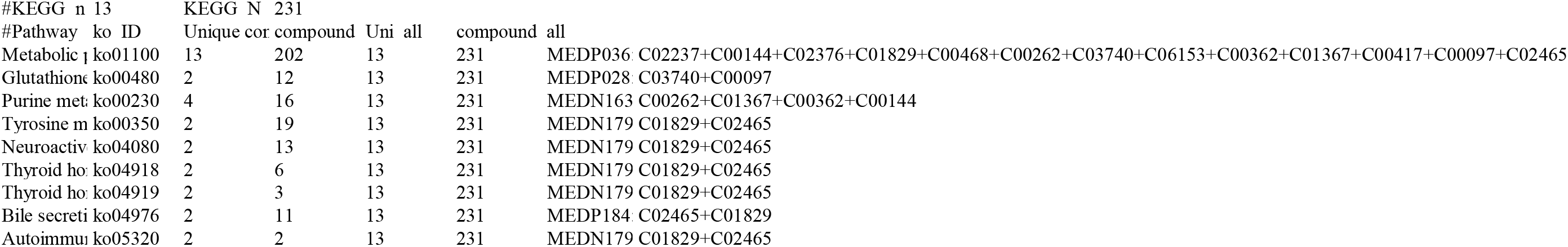
KEGG enrichment analysis of DEMs unique to F vs H.

**Table S8.**
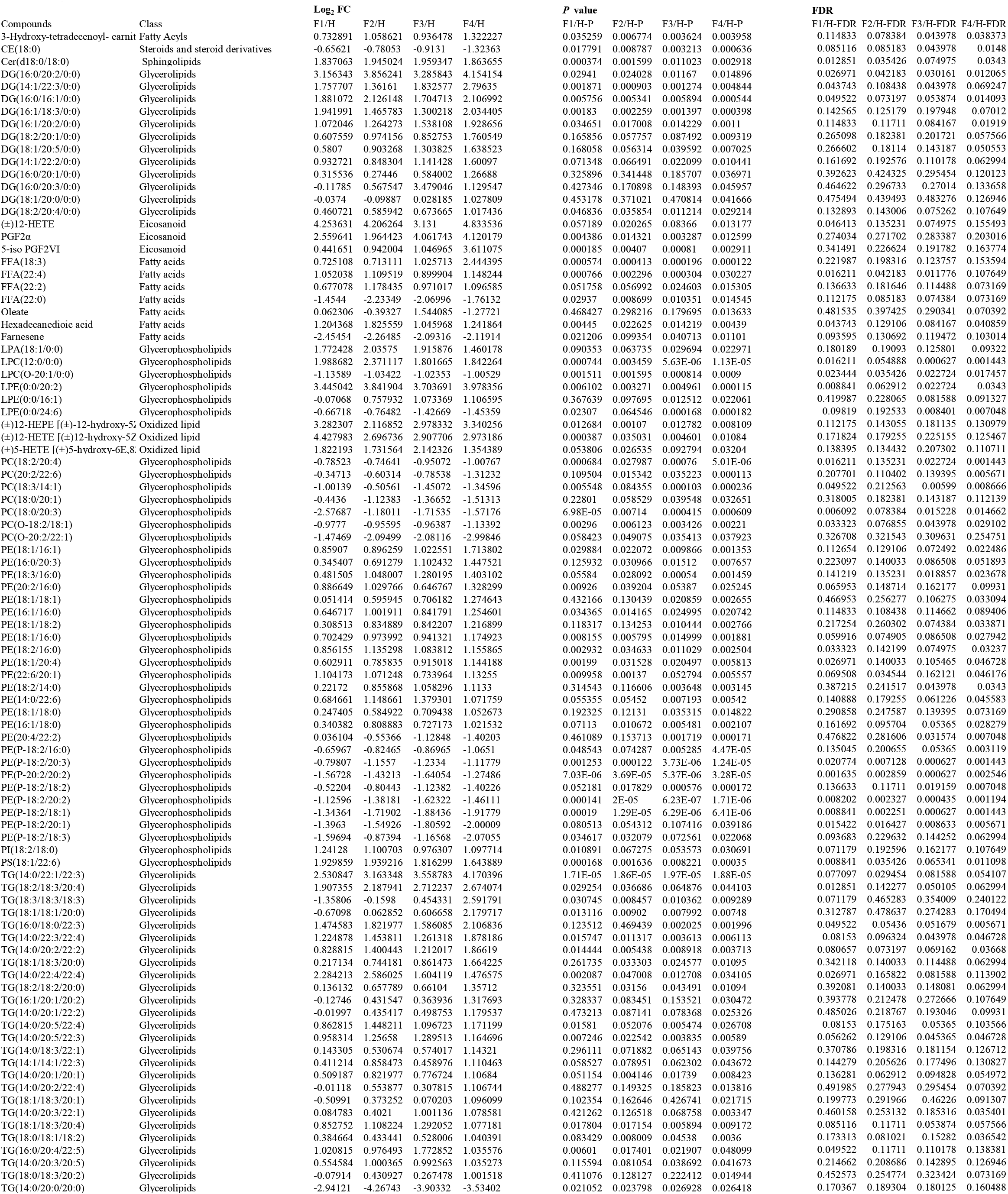
Lipidomic data of F vs H.

**Table S9.**
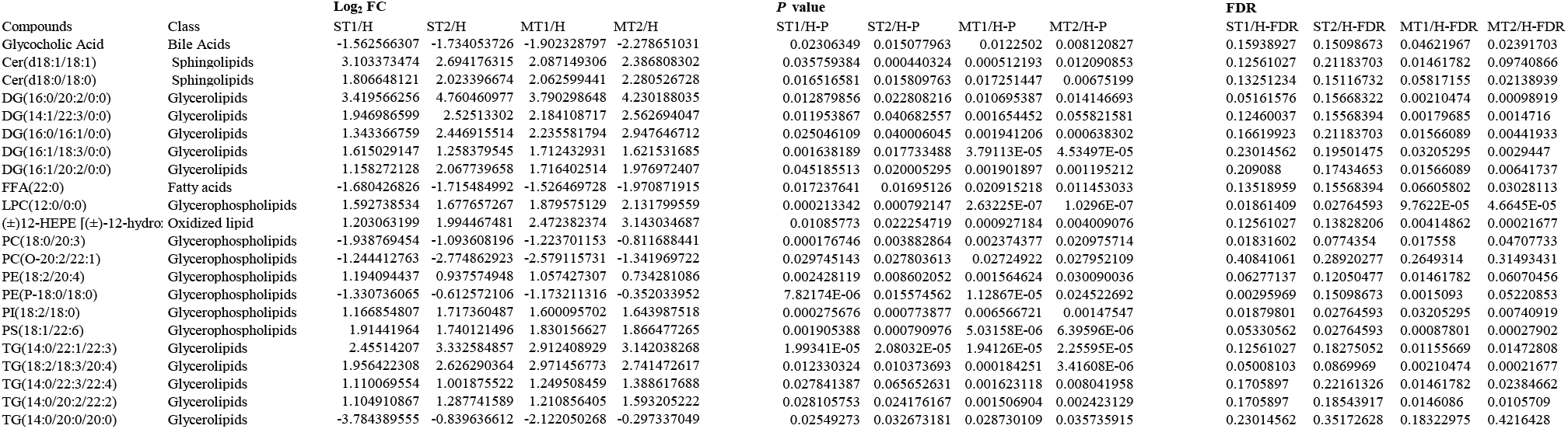
Lipidomic data of S vs H and M vs H.

**Table S10.**
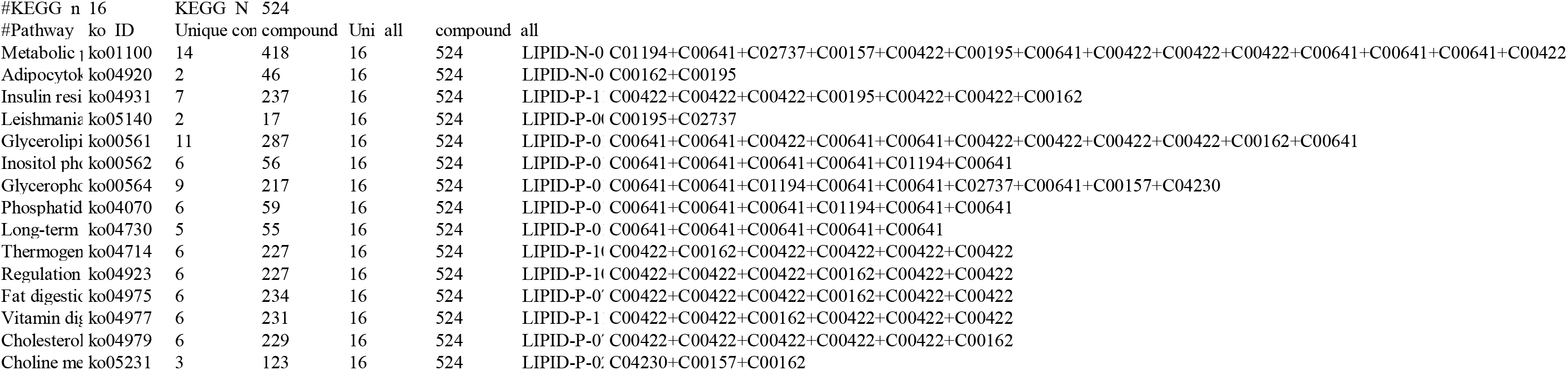
KEGG enrichment analysis of DEIs shared by F vs H, S vs H and M vs H.

**Table S11.**
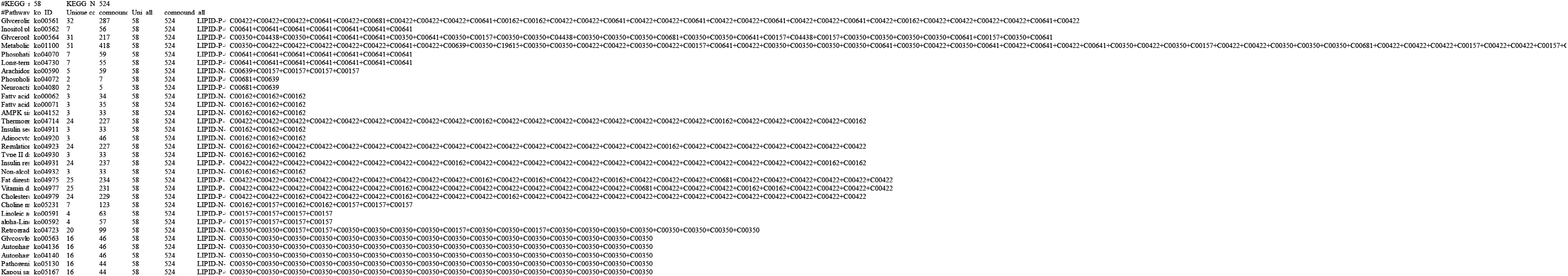
KEGG enrichment analysis of DEIs unique to F vs H.

**Table S12.**
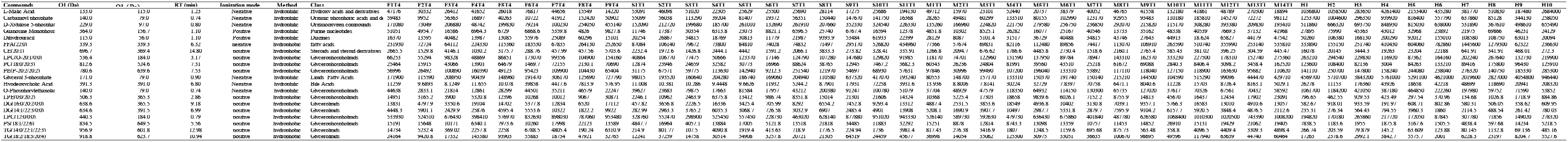
Normalized expression levels of potential biomarkers.

**Table S13.**
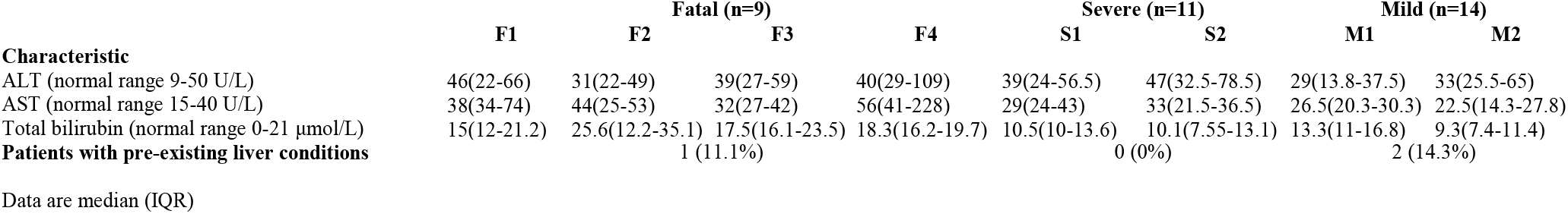
Clinical characteristics of COVID-19 patients in this study.

**Table S14.**
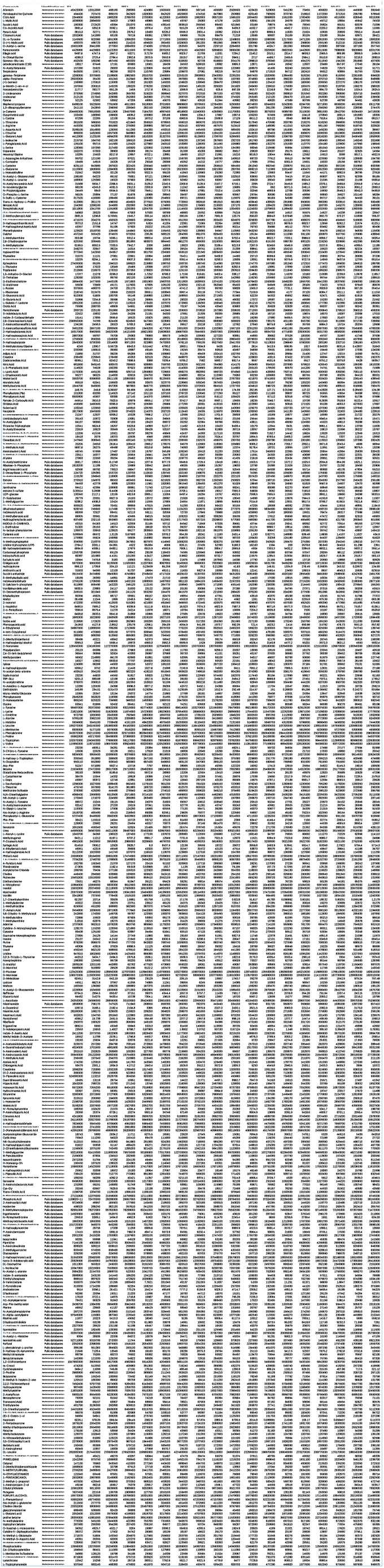
Metabolite identification methods and datasets.

**Table S15.**
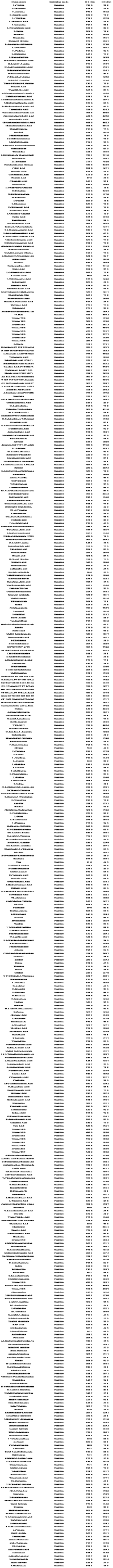
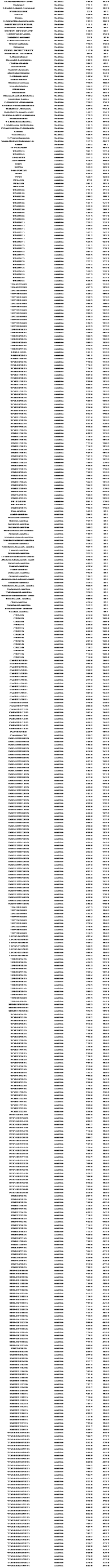
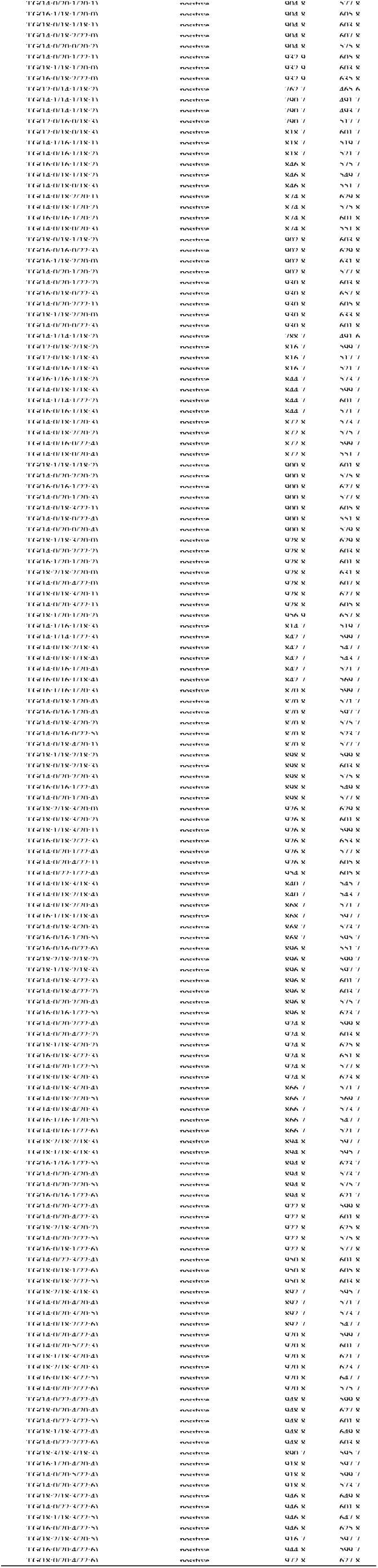
The ionization modes and ion pairs of the metabolites.

